# Operational Heterogeneity of Capillary Refill Time Across Two Public ICU Databases: A Methodological Analysis

**DOI:** 10.64898/2026.09.29.26364303

**Authors:** Xue Xia, Zhiyong Xie

## Abstract

**Background:** Capillary refill time (CRT) is an increasingly endorsed bedside perfusion marker in septic shock. Publicly available intensive care unit (ICU) databases such as MIMIC-IV and eICU-CRD enable large-scale multi-database research, but the cross-database comparability of CRT operationalization has not been systematically examined.

**Methods:** We conducted a retrospective, descriptive, cross-database methodological analysis of adult ICU stays in MIMIC-IV v3.1 (2008–2022) and eICU-CRD v2.0 (2014–2015). CRT availability, definitional thresholds, recording patterns and time windows, bilateral measurement consistency, and selection bias in the CRT-documented population were characterized descriptively.

**Results:** Among 94,458 MIMIC-IV and 200,234 eICU-CRD adult ICU stays, CRT availability differed markedly: 79.85% of MIMIC-IV stays had at least one CRT record within the first 24 hours, versus 6.35% of eICU-CRD stays across the entire ICU stay, a 73.5-percentage-point gap. The abnormality threshold was 3 seconds in MIMIC-IV and 2 seconds in eICU-CRD. Among CRT-available MIMIC-IV stays, the mean number of CRT records in the first 24 hours was 7.0 (median 6, interquartile range (IQR) 4–10); 99.6% had bilateral records, with 99.4% left–right agreement (Cohen’s kappa 0.914). The temporal distribution of CRT records was strikingly similar between databases: only 28.7% of MIMIC-IV and 29.1% of eICU-CRD records occurred within 24 hours of ICU admission (including pre-admission records), with roughly 71% in both databases recorded later in the stay. In eICU-CRD, the CRT-documented population had higher illness severity than the undocumented population (Acute Physiology and Chronic Health Evaluation (APACHE) IV 60.0 versus 54.4; standardized mean difference 0.21), consistent with selection by indication. Structured peripheral perfusion index fields were unavailable in both databases.

**Conclusions:** CRT operationalization differs fundamentally across MIMIC-IV and eICU-CRD in availability, definitional thresholds, recording patterns, bilateral structure, and patient selection. Cross-database CRT research should explicitly declare definitional heterogeneity, avoid pooling across thresholds, and treat sparsely documented subsets as selected populations. Selection bias was evident in both databases, in opposite directions. Severity-score-based comparison was possible only in eICU-CRD. Standardized common data elements for bedside perfusion assessment are needed to make cross-database CRT research valid by design.

## 1. Background

Capillary refill time (CRT), defined as the time required for skin color to return to baseline after blanching pressure is applied to a distal capillary bed, is a bedside perfusion assessment increasingly endorsed as an adjunct marker in septic shock resuscitation. The Surviving Sepsis Campaign first suggested using CRT to guide resuscitation as an adjunct to other measures of perfusion in 2021, a recommendation retained in the 2026 update (conditional recommendation, low certainty of evidence) [1]. In late 2025, the ANDROMEDA-SHOCK-2 multicenter randomized trial (n = 1,467; 86 centers in 19 countries) demonstrated that a personalized hemodynamic resuscitation protocol targeting CRT normalization was superior to usual care on a hierarchical composite of mortality, duration of vital support, and hospital length of stay (win ratio 1.16, 95% confidence interval (CI) 1.02–1.33). The benefit was driven mainly by shorter duration of vital support; 28-day mortality did not differ significantly (26.5% vs 26.6%) [2]. These findings have renewed interest in CRT as both a therapeutic target and a prognostic predictor in critical illness, with its prognostic value demonstrated in large prospective studies [3] and confirmed in a recent systematic review and meta-analysis [4].

Publicly available ICU databases, including the single-center Medical Information Mart for Intensive Care IV (MIMIC-IV) [5] and the multi-center eICU Collaborative Research Database (eICU-CRD) [6], have become foundational resources for critical care research. External validation applies a prediction model developed in one setting to an independent dataset and is a rigorous and widely endorsed approach for assessing the generalizability of clinical prediction models [7]. However, the validity of cross-database comparisons depends critically on consistent operational definitions of the variables being compared. For bedside measurements such as CRT, which are recorded as part of routine clinical documentation rather than under a standardized research protocol, operational definitions may differ between databases in ways that are difficult to detect without systematic examination. To our knowledge, no prior study has systematically compared the operationalization of CRT across public ICU databases.

We characterized CRT operationalization in MIMIC-IV v3.1 and eICU-CRD v2.0 along five dimensions: data availability, definitional thresholds, recording patterns and time windows, bilateral measurement consistency, and selection bias in the CRT-documented population. Our objective was methodological rather than clinical: to provide an empirical foundation for cross-database CRT research and to identify pitfalls that may compromise the validity of multi-database prediction studies. These pitfalls bear directly on how cross-database CRT prediction models should be designed and interpreted.

## 2. Methods

### 2.1 Study design and setting

We conducted a retrospective, descriptive, cross-database methodological analysis using two publicly available ICU databases: MIMIC-IV v3.1 [5] and eICU-CRD v2.0 [6]. MIMIC-IV contains de-identified health data from Beth Israel Deaconess Medical Center (Boston, MA), encompassing comprehensive electronic health record (EHR) data for patients admitted between 2008 and 2022. eICU-CRD is a multi-center database containing ICU data from over 200 hospitals across the United States, collected between 2014 and 2015. Both databases were accessed through PhysioNet [8] and extracted on 2026-09-02. Source data were loaded into a local DuckDB analysis environment [9] with SHA256 verification after download. This study is a secondary analysis of existing, de-identified public datasets; the oversight arrangements for the source databases are set out in the Declarations. Reporting followed STROBE [10] and RECORD [11] guidelines for observational studies conducted using routinely collected health data (Supplementary files 1 and 2).

### 2.2 Participants

The analysis population comprised all adult ICU stays in each database. In MIMIC-IV, 0 of 364,627 patients in the patients table had anchor_age < 18 years, and all 94,458 ICU stays were retained. In eICU-CRD, the age field is stored as a string with ages above 89 top-coded as “> 89”; we mapped these to 90 for age filtering and descriptive statistics. Of 200,859 ICU stays, 625 (0.31%) were excluded because age was below 18 years (n = 530) or not recorded (n = 95), yielding 200,234 adult ICU stays. No further inclusion or exclusion criteria were applied at cohort selection.

### 2.3 Variables: CRT definitions

#### 2.3.1 MIMIC-IV CRT

In MIMIC-IV, CRT was identified through a systematic search of the d_items dictionary for labels or abbreviations containing “capillary” or “refill”, which returned two itemids, both charted in chartevents: 223951 (R) and 224308 (L) (Supplementary file 3). The search returned no other candidate items. Both itemids recorded only two distinct values, “Normal <3 Seconds” and “Abnormal >3 Seconds”; the operative threshold was therefore 3 seconds.

#### 2.3.2 eICU-CRD CRT

In eICU-CRD, CRT was identified in the nurseAssessment table; an initial search of nurseCharting returned no CRT records. Under the “Capillary Refill” cellattribute, state and site values are stored in a single cellattributevalue column, with the state and site of the same assessment appearing as separate rows. Values are classified by semantic content as either state (normal, < 2 seconds, > 2 seconds) or site (hands, feet). For comparability, the state values were collapsed to a binary variable at a 2-second threshold: normal and < 2 seconds were treated as normal, and > 2 seconds as abnormal. Co-occurrence of hand and foot site rows was examined at the stay level; stays were classified as both-sites, hands-only, or feet-only (Supplementary file 4).

### 2.4 Supplementary perfusion variables

Supplementary perfusion-related variables were identified in four eICU-CRD physicalExam pathways ending in “Extremities/Perfusion/adequate”, “Extremities/Perfusion/extremities cool”, “Extremities/Perfusion/extremities cold/dusky”, and “Extremities/Perfusion/extremities flushed” (Table 5). Structured peripheral perfusion index (PPI) fields were unavailable in both databases: no fields matching “perfusion index” or “PPI” were found in the MIMIC-IV d_items dictionary or the eICU-CRD physicalExam table.

### 2.5 Time window characterization

In MIMIC-IV, CRT record times were expressed as hours from ICU admission (charttime − icustays.intime). In eICU-CRD, the native nurseassessoffset (minutes from unit admission) was converted to hours; the 6-, 12-, and 24-hour window boundaries correspond to offsets of 360, 720, and 1440 minutes. Records falling before ICU admission were classified as pre-admission; all other records were assigned to 0–6h, 6–12h, 12–24h, or >24h windows (half-open intervals, lower bound inclusive). The distribution of CRT records across these windows was compared between databases to assess temporal alignment of CRT recording. The present manuscript reports the preregistered methodological aim in full; all analyses reported herein are descriptive and methodological in nature, consistent with the feasibility-probe scope declared at registration (Supplementary file 5).

### 2.6 Selection bias quantification

To evaluate selection bias in CRT documentation, we compared baseline characteristics and outcomes between CRT-available and CRT-unavailable adult ICU stays in both databases (MIMIC-IV: CRT recorded within the first 24 hours; eICU-CRD: CRT recorded across the entire ICU stay). In eICU-CRD, characteristics compared were age, sex, APACHE IV score, acute physiology score, predicted ICU mortality, and actual hospital mortality. In MIMIC-IV, where standardized severity scores were unavailable, we compared age, sex, hospital mortality (from admissions.deathtime), and ICU length of stay as proxy measures for illness severity. Age in MIMIC-IV was calculated as anchor_age plus the difference between the year of ICU admission and anchor_year, per MIMIC-IV documentation; in eICU-CRD, age was taken from the admission record (values > 89 mapped to 90, as described in Section 2.2).

### 2.7 Statistical analysis

All analyses were descriptive. Continuous measures were summarized using means and standard deviations, and categorical measures using counts and percentages. Bilateral consistency in MIMIC-IV was assessed using percent agreement and Cohen’s kappa between left-hand and right-hand measurements. Left- and right-hand records were paired at the stay level using the most recent record of each side within the first 24 hours; both sides were required to have at least one such record. Agreement was defined as the proportion of bilateral stays where both sides yielded the same binary classification. For eICU-CRD, hand and foot records were paired at the stay level using the most recent assessment per site; concordance was defined as the proportion of paired stays with valid state values at both sites and the same binary classification (Supplementary file 4). Because this was an operational heterogeneity study, no multivariable modeling, causal inference, or hypothesis testing was performed. Between-group differences in continuous measures were also expressed as standardized mean differences (SMD), calculated as the difference in group means divided by the pooled standard deviation (sample-size-weighted). No imputation was performed; variables without recorded values were reported as unavailable. All analyses were performed using DuckDB (v1.5.5) on a local workstation.

### 2.8 Use of AI-assisted technologies in data analysis

During the preparation of this study, the authors used DeepSeek-V4 Pro (DeepSeek; accessed August–September 2026) to assist in drafting SQL extraction queries and Python data-processing scripts for the local DuckDB analysis environment. All AI-assisted code and analytic outputs were independently reviewed and validated by the authors against SHA256-verified source data, by comparing query outputs against source record counts and by manual review of SQL logic. The authors take full responsibility for the integrity and validity of all code and results presented in this manuscript.

## 3. Results

### 3.1 Study databases

The analysis included 94,458 adult ICU stays from MIMIC-IV and 200,234 adult ICU stays from eICU-CRD (Figure 1). MIMIC-IV provided a relatively high-coverage environment for structured CRT documentation, whereas eICU-CRD provided broader hospital coverage but substantially sparser CRT availability.

**Figure 1.**
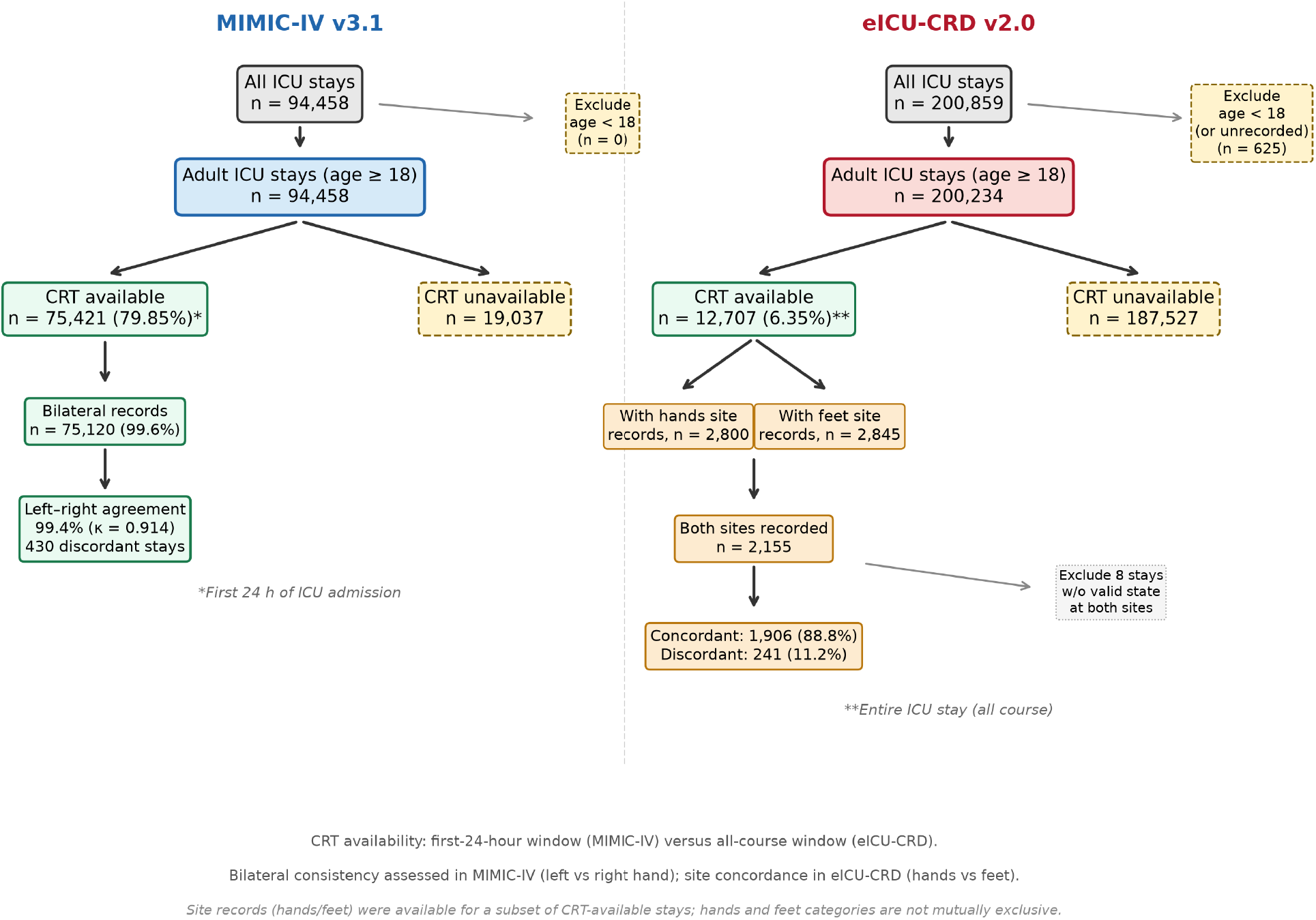
ICU stay flow diagram. MIMIC-IV (left): all adult ICU stays with CRT availability calculated within the first 24 hours of ICU admission. eICU-CRD (right): adult ICU stays after excluding patients aged less than 18 years, with CRT availability calculated across the entire ICU stay. Bilateral consistency was assessed in MIMIC-IV (left hand versus right hand); site concordance was assessed in eICU-CRD (hands versus feet).

### 3.2 CRT availability

CRT availability was markedly higher in MIMIC-IV than in eICU-CRD (Table 1, Figure 2A). In MIMIC-IV, 75,421 of 94,458 adult stays (79.85%) had at least one CRT record in the first 24 hours of ICU admission. In eICU-CRD, 12,707 of 200,234 adult stays (6.35%) had at least one CRT record across the entire ICU stay (all-course). This absolute difference of approximately 73.5 percentage points indicates that CRT availability is the dominant source of cross-database heterogeneity. Across the entire ICU stay, MIMIC-IV CRT availability was 81,336 of 94,458 stays (86.1%), so the 73.5-percentage-point gap is, if anything, conservative relative to an all-course-versus-all-course comparison.

**Table 1.** Operational comparison of CRT definitions across MIMIC-IV and eICU-CRD.

| Domain | MIMIC-IV v3.1 | eICU-CRD v2.0 |
| --- | --- | --- |
| Source table | chartevents | nurseAssessment |
| Item identifier | itemid 223951 (R), 224308 (L) | cellattribute "Capillary Refill" |
| Threshold | 3 seconds | 2 seconds |
| Value format | Binary: Normal / Abnormal | Multi-state: normal, <2s, >2s + site (hands/feet) |
| Availability (n / N) | 75,421 / 94,458 (79.85%) | 12,707 / 200,234 (6.35%) |
| Mean records / stay | 7.0 (median 6, IQR 4–10) | 27.5 (median 13, IQR 6–29) |
| Bilateral available | 99.6% | Not applicable (hands/feet sites) |
| Left–right agreement | 99.4% ($\kappa = 0.914$) | Not applicable |

MIMIC-IV: first-24-hour window. eICU-CRD: all-course (stay-level availability calculated over full ICU stay; record-level time windows are reported in Table 3); includes state and site records (271,657 state + 78,068 site = 349,725 records across 12,707 stays). eICU total adult stays: 200,234.

**Figure 2.**
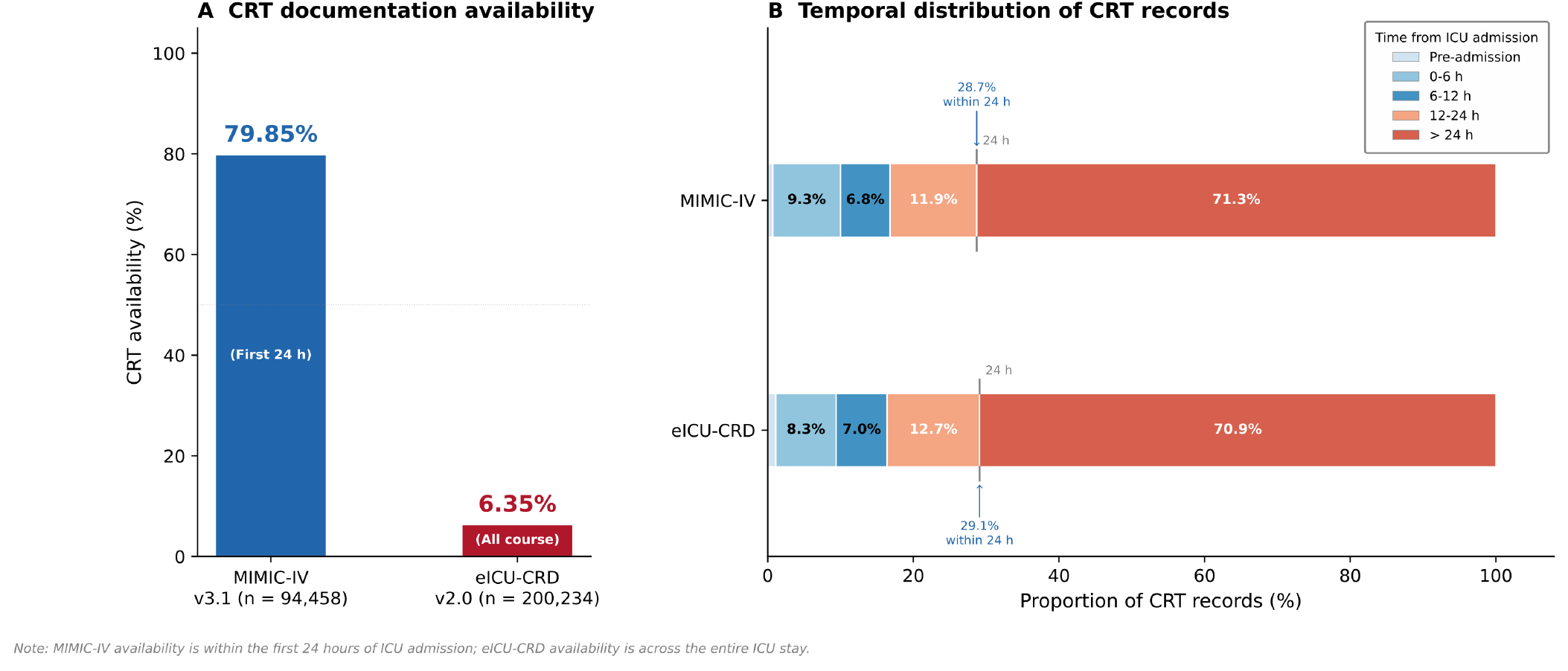
Cross-database comparison of CRT documentation. **A**, CRT availability: 79.85% in MIMIC-IV (first-24-hour window) versus 6.35% in eICU-CRD (all-course). **B**, Temporal distribution of CRT records from ICU admission; the majority of records in both databases fall after 24 hours (71.3% in MIMIC-IV, 70.9% in eICU-CRD). Tick marks indicate the 24-hour boundary for each database (MIMIC-IV 28.7%, eICU-CRD 29.1%, including pre-admission records). Pre-admission segments (MIMIC-IV 0.7%, eICU-CRD 1.1%) are too small to label.

### 3.3 Threshold definitions and value structure

The two databases operationalized CRT using different thresholds and storage structures. MIMIC-IV used a 3-second cutoff stored as binary item values (“Normal <3 Seconds” or “Abnormal >3 Seconds”) in the chartevents table. eICU-CRD used a 2-second conceptual cutoff embedded within the nurseAssessment table, with mixed state and site attributes. The state values included “normal,” “< 2 seconds,” and “> 2 seconds,” while the site values indicated “hands” or “feet.” Because eICU-CRD contains both “normal” and “< 2 seconds” as distinct state values, the semantic equivalence of these categories cannot be confirmed from the stored data alone. For the purposes of this analysis, both “normal” and “< 2 seconds” were combined into a single “normal” category, and “> 2 seconds” was classified as “abnormal.”

The eICU-CRD value distribution across the 349,725 CRT-related records in the adult cohort was as follows: “normal” 51.5% (180,145 records, 10,914 stays), “< 2 seconds” 13.4% (46,975 records, 4,191 stays), “> 2 seconds” 12.7% (44,537 records, 3,168 stays), “hands” 10.6% (37,009 records, 2,799 stays), and “feet” 11.7% (41,059 records, 2,845 stays). The co-occurrence of hands and feet site records was examined: 2,155 stays had both sites recorded, while 644 had hands only and 690 had feet only.

State-level stay counts are not mutually exclusive; a single stay may contribute to multiple categories across time. The total number of stays with any state-level CRT record (12,707) is therefore less than the sum of category-specific stay counts.

### 3.4 Recording patterns

Among adult CRT-available MIMIC-IV stays, the mean number of CRT records per stay was 7.0 (median 6, interquartile range 4–10; Table 2).

**Table 2.**
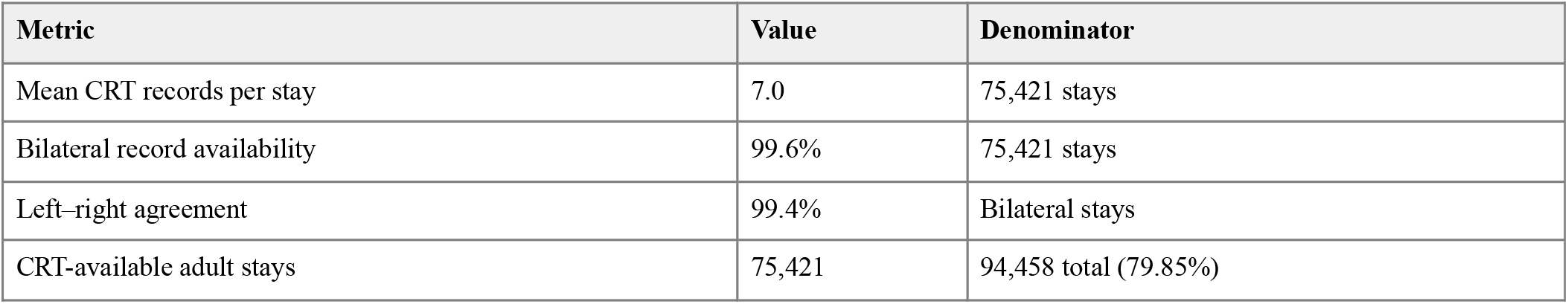
CRT recording patterns in MIMIC-IV among CRT-available adult stays.

| Metric | Value | Denominator |
| --- | --- | --- |
| Mean CRT records per stay | 7.0 | 75,421 stays |
| Bilateral record availability | 99.6% | 75,421 stays |
| Left–right agreement | 99.4% | Bilateral stays |
| CRT-available adult stays | 75,421 | 94,458 total (79.85%) |

### 3.5 Recording time windows

The temporal distribution of CRT records was strikingly similar across databases (Table 3, Figure 2B). In MIMIC-IV, 28.7% of CRT records fell within the first 24 hours of ICU admission (0.7% pre-admission, 9.3% at 0–6h, 6.8% at 6– 12h, 11.9% at 12–24h), while 71.3% occurred after 24 hours. In eICU-CRD, 29.1% of CRT records fell within the first 24 hours (1.1% pre-admission, 8.3% at 0–6h, 7.0% at 6–12h, 12.7% at 12–24h), with 70.9% occurring after 24 hours. The 24-hour cutpoint corresponds to the standard predictor ascertainment window in ICU outcome prediction studies.

**Table 3.** Distribution of CRT records by time from ICU admission.

| Time window | MIMIC Records | MIMIC % | eICU Records | eICU % |
| --- | --- | --- | --- | --- |
| Pre-admit | 12,871 | 0.7 | 3,076 | 1.1 |
| 0–6h | 176,199 | 9.3 | 22,502 | 8.3 |
| 6–12h | 128,512 | 6.8 | 19,092 | 7.0 |
| 12–24h | 225,200 | 11.9 | 34,495 | 12.7 |
| >24h | 1,349,369 | 71.3 | 192,492 | 70.9 |

### 3.6 Selection bias: CRT-available vs CRT-unavailable populations

Selection bias was examined in both databases. In MIMIC-IV, CRT-available stays (n = 75,421) differed from CRT-unavailable stays (n = 19,037) in the opposite direction from eICU-CRD: CRT-available patients were younger (mean age 64.6 vs 65.5 years), had a higher proportion of male patients (56.4% vs 54.2%), lower hospital mortality (11.3% vs 14.7%), and longer ICU stays (mean 3.8 vs 3.0 days).

In eICU-CRD, to evaluate whether the CRT-available population represents a clinically distinct subgroup, baseline characteristics and outcomes were compared between CRT-available and CRT-unavailable adult ICU stays (Table 4). Although age and sex were similar between groups (mean age 63.2 (SD 16.9) and 63.2 (SD 17.0) years, respectively; male 55.1% vs 53.9%), illness severity markers differed substantially. APACHE IV scores were higher in the CRT-available group (60.0 vs 54.4), as were acute physiology scores (48.5 vs 42.7) and predicted ICU mortality (6.0% vs 4.9%). Actual hospital mortality was 9.8% in the CRT-available group versus 8.9% in the CRT-unavailable group.

**Table 4.**
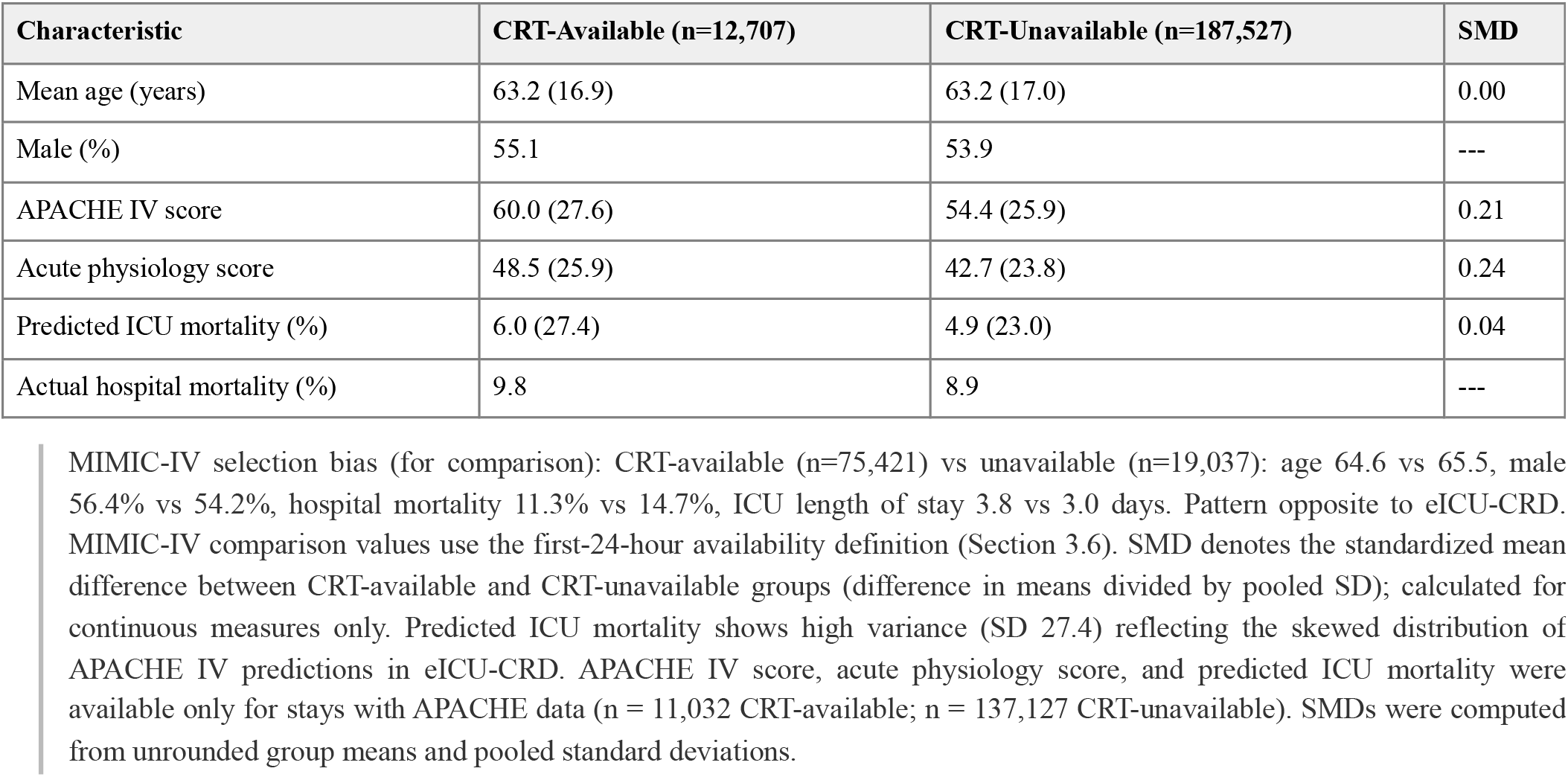
Baseline characteristics and outcomes of eICU-CRD adult ICU stays by CRT availability (continuous measures as mean (SD); hospital mortality as %)

These differences indicate that eICU CRT documentation is preferentially triggered for sicker patients, consistent with selection by indication. When only 6.35% of stays have CRT recorded, the recorded population is unlikely to be representative of all ICU patients. This has important implications for cross-database prediction: the eICU “CRT-available” population differs meaningfully from the MIMIC population, where CRT coverage approaches 80%.

### 3.7 Bilateral consistency

In MIMIC-IV, bilateral record availability was 99.6% among adult CRT-available stays, and left–right agreement was 99.4% (Cohen’s kappa = 0.914), reflecting near-perfect concordance even after accounting for the high prevalence of normal values (96.3% of records; abnormal prevalence 3.7%). The 430 discordant stays (0.6%) comprised 238 with right-normal/left-abnormal and 192 with right-abnormal/left-normal patterns. Left- and right-hand measurements were recorded at the same timestamp in 98.2% of bilateral stays (mean time difference 0.12 hours), confirming that paired comparisons reflect concurrent assessments rather than temporally separated observations. The low abnormal prevalence means the high agreement rate should be interpreted in context.

### 3.8 Supplementary perfusion variables

eICU-CRD contained supplementary perfusion-related observations in the physicalExam table (Table 5), including perfusion adequate (51,046 records across 9,724 stays), extremities cool (5,102 records across 1,097 stays), extremities cold/dusky (717 records across 188 stays), and extremities flushed (158 records across 48 stays). Among 2,155 eICU-CRD stays with both hands and feet CRT records (out of 2,799 stays with hands records and 2,845 with feet records), concordance at the most recent paired assessment was observed in 1,906 of 2,147 stays with valid state values for both sites (88.8%); the 241 discordant stays (11.2%) showed a strong directional asymmetry, with hands recorded as normal and feet as abnormal in 198 stays (9.2%) versus the reverse in only 43 stays (2.0%). This hands/feet dimension is distinct from the left/right bilateral consistency assessed in MIMIC-IV and cannot be directly compared across databases.

**Table 5.**
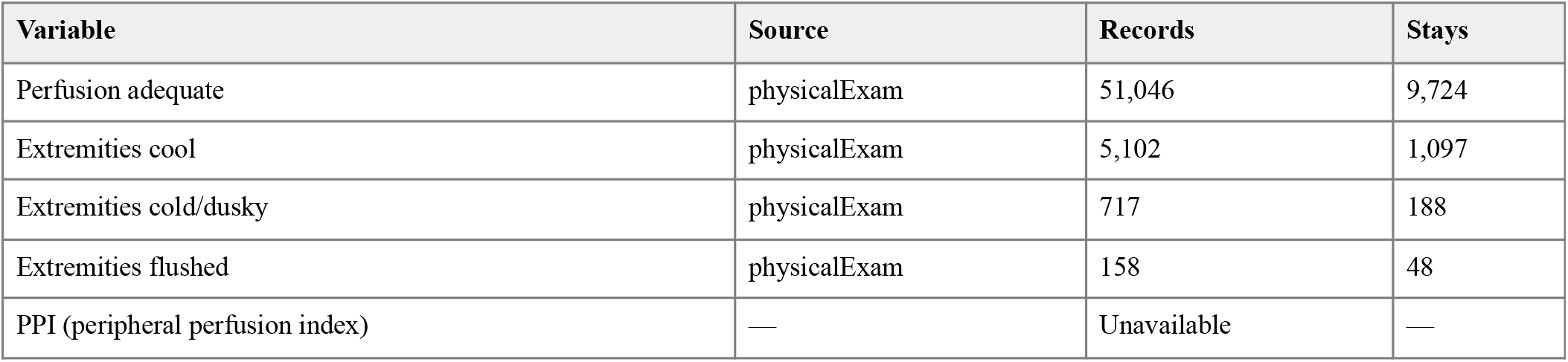
Supplementary perfusion variables in eICU-CRD physicalExam table.

### 3.9 Cross-database definitional mapping

The mapping between databases highlighted structural differences beyond the numeric threshold. In MIMIC-IV, CRT was a discrete bedside observation item with clear left/right identifiers (itemid 223951 and 224308) and a consistent binary value format. In eICU-CRD, CRT was embedded within a broader nursing assessment structure with cellattribute-based storage and mixed state/site attributes. These differences indicate that cross-database CRT comparisons require explicit mapping rules rather than assumptions of direct equivalence.

The key mapping challenges include: (1) the threshold discrepancy (3 seconds vs 2 seconds), which means the same patient could be classified differently under each definition; (2) the semantic ambiguity of eICU-CRD state values (“normal” vs “< 2 seconds”), which cannot be resolved without external documentation; and (3) the site attribute structure in eICU-CRD (hands/feet recorded as separate rows), which has no direct MIMIC-IV counterpart. These challenges underscore the need for explicit harmonization protocols in any future cross-database CRT prediction study.

## 4. Discussion

### 4.1 Summary of principal findings

In this cross-database methodological analysis of 294,692 adult ICU stays, we found that capillary refill time (CRT) is operationalized in fundamentally different ways in MIMIC-IV and eICU-CRD. The dominant source of heterogeneity was availability: four of five MIMIC-IV stays had at least one CRT record, compared with fewer than one in fifteen in eICU-CRD. The databases also applied different abnormality thresholds (3 vs 2 seconds) and stored CRT in structurally different formats, yet exhibited a strikingly similar temporal distribution of records (∼29% within the first 24 hours in both). Bilateral consistency in MIMIC-IV was high, whereas in eICU-CRD the recording structure (hands/feet sites rather than left/right pairs) precluded an equivalent analysis. Finally, the selection patterns differed between databases: in eICU-CRD, the CRT-documented population was a severity-selected subgroup with higher illness severity and mortality, while in MIMIC-IV, CRT documentation reflected routine practice in longer-staying patients with lower mortality.

### 4.2 Interpretation by dimension

#### Availability

The 73.5-percentage-point coverage gap (79.85% vs 6.35%; approximately 12.6-fold) indicates that CRT documentation is near-routine in MIMIC-IV but exceptional in eICU-CRD. Importantly, this is a problem of availability rather than missingness: within CRT-available stays, records are abundant. The gap therefore most plausibly reflects differences in nursing documentation practice across the contributing hospitals rather than data capture failure, an interpretation supported by the richness of the eICU-CRD records where they do exist (349,725 records across five state and site values). For researchers, the practical consequence is that any eICU-CRD analysis conditioned on CRT availability discards 93.7% of the population before analysis begins, and does so non-randomly (see Selection bias, below).

#### Thresholds

MIMIC-IV encodes CRT as a binary value against a 3-second cutoff, whereas eICU-CRD embeds a 2-second conceptual cutoff within a multi-state nursing assessment field. A patient whose true CRT lies between 2 and 3 seconds would be classified as normal in MIMIC-IV but abnormal in eICU-CRD, a directional and systematic misclassification that no analytic harmonization can reverse, because neither database stores the underlying measurement in seconds. Pre-classified text values cannot be re-derived against an alternative threshold. Notably, the 3-second cutoff aligns MIMIC-IV with the standardized definition used in the ANDROMEDA-SHOCK trials [2, 12, 13], making it the more natural choice for studies seeking trial-comparable phenotypes. The practical impact of this threshold discordance is substantial: the abnormal CRT prevalence was 3.7% of MIMIC-IV records versus 16.4% of eICU-CRD records carrying a CRT state value (12.7% of all CRT-related records, including site-only rows), roughly one-quarter, and the resulting case mix differs markedly between databases.

#### Recording patterns and time windows

The repeated-measurement environment in MIMIC-IV (mean 7.0 records per stay) supports worst-value summary rules and temporal trend assessment, whereas reliance on a single first value would discard most available information. However, temporal alignment across databases was in fact remarkably consistent: 28.7% of MIMIC-IV records and 29.1% of eICU-CRD records fell within the first 24 hours of ICU admission, the window most commonly used for predictor ascertainment in outcome prediction studies (Table 3, Figure 2B). In both databases, roughly 71% of records occurred after 24 hours, suggesting that CRT documentation reflects ongoing reassessment throughout the ICU stay rather than admission-time screening. A small fraction of records in both databases carried pre-admission timestamps (0.7% in MIMIC-IV, 1.1% in eICU-CRD; medians 0.6 and 0.3 hours before admission, respectively), a documentation-timing artifact that cross-database studies should anticipate and handle explicitly. Although most pre-admission records clustered within approximately one hour of ICU admission, both databases contained small long tails extending far beyond 48 hours (MIMIC-IV: 735 records more than 48 hours before admission, including 358 more than 7 days and 46 more than 30 days; eICU-CRD: 91 records more than 48 hours). The most extreme MIMIC-IV record preceded ICU admission by approximately one year, consistent with a charted-year error rather than a genuine pre-admission measurement.

#### Bilateral consistency

In MIMIC-IV, 99.6% of CRT-available stays had bilateral records and left–right agreement was 99.4% (Cohen’s kappa = 0.914), supporting simplified operational rules such as single-side abstraction or worst-value strategies; the 0.6% discordant cases (430 stays) are unlikely to materially affect binary classification. No equivalent statement is possible for eICU-CRD: its site attributes (hands/feet) describe distal anatomical sites rather than lateral pairs, addressing a different physiological question (an upper-versus lower-extremity perfusion gradient) with no direct MIMIC-IV counterpart, and only 2,155 stays had both sites recorded. Among them, hands and feet agreed in 88.8% of stays with valid state values at both sites, and the discordance was strongly directional (hands normal with feet abnormal in 198 stays versus the reverse in 43), consistent with a distal-to-proximal perfusion gradient rather than random disagreement. Bilateral consistency is therefore effectively a MIMIC-IV-only dimension, and any cross-database “bilateral” comparison would be a category error.

#### Selection bias

The direction of selection differed between databases. In eICU-CRD, the CRT-available population was more severely ill (APACHE IV 60.0 vs 54.4; SMD 0.21; Table 4), consistent with severity-triggered documentation: clinicians chart CRT when they have reason to assess peripheral perfusion, as expected under informed presence bias [14, 15]. Actual hospital mortality was only modestly higher (9.8% vs 8.9%). In MIMIC-IV, the pattern was reversed: CRT-available patients were younger (64.6 vs 65.5 years), had lower hospital mortality (11.3% vs 14.7%), and longer ICU stays (3.8 vs 3.0 days) than CRT-unavailable patients, suggesting that CRT documentation in MIMIC-IV reflects routine nursing practice in longer-staying patients rather than acute severity triggers. This asymmetry has direct implications for cross-database prediction research: the eICU CRT-available subset estimates model performance in a severity-shifted spectrum [16–18], while the MIMIC CRT-available subset captures a broader, less selected population. Apparent performance differences between databases may partly reflect these opposing selection patterns rather than true transportability failure.

### 4.3 Comparison with previous work

To our knowledge, no prior study has systematically quantified the operationalization of CRT across public ICU databases. This study extends prior methodological work in three directions. First, the measurement-standardization literature has long documented that CRT values depend on procedural and environmental factors (technique, site, skin and room temperature, lighting, and patient characteristics), motivating standardized protocols [13, 19]. Our findings show that even after measurement, the *recorded representation* of CRT inherits a second layer of heterogeneity (thresholds, storage structures) that persists regardless of bedside technique; the 99.4% left–right agreement (Cohen’s kappa = 0.914) we observed reflects documentation consistency within a recording workflow, not measurement reliability, and should not be interpreted as evidence that CRT measurement itself is standardized [20]. The largest interobserver study found only moderate agreement (κ = 0.56) [20].

Second, the EHR data-quality literature has established frameworks [21–23], and cross-site validation has documented transportability challenges [24]. Our results provide a concrete, quantified case study for known barriers to prediction model transportability. Riley and colleagues identified differential measurement, different measurement time points, and different cut-offs as causes of heterogeneity in predictor effects across settings [7]; CRT across MIMIC-IV and eICU-CRD exhibits all three simultaneously, plus severity-selected availability. Empirical transportability studies have similarly identified EHR configuration and data warehouse heterogeneity as barriers to cross-site validation. The magnitude we document, a 12.6-fold availability ratio with a discordant abnormality threshold, is at the extreme end of what this literature anticipates for routinely documented variables.

Third, the contrast with trial-grade measurement is instructive. The ANDROMEDA-SHOCK trials measured CRT with a standardized protocol (blanching pressure applied with a glass microscope slide to the right index finger for 10 seconds, timed with a chronometer, abnormal defined as > 3 seconds) [2, 12, 13]. Neither database records measurement conditions, so researchers cannot determine how closely routine documentation approximates this protocol; database-derived CRT phenotypes should therefore be described as “documented CRT” rather than assumed equivalent to protocolized CRT. CRT is also known to lack structured representation in electronic health records [22], and peripheral perfusion assessment in critical illness may benefit from more standardized measurement approaches [25].

### 4.4 Implications and recommendations

For researchers planning cross-database CRT studies, including prediction models of mortality or resuscitation response, we offer five recommendations:

1. **Report availability as a data-quality measure**. CRT coverage (79.85% vs 6.35% here) should be stated explicitly in any database study of routinely documented bedside variables, alongside conventional missingness statistics [26].
2. **Document threshold definitions explicitly, and do not pool across discordant thresholds**. When thresholds differ (3 vs 2 seconds), pooled analysis without recalibration embeds directional misclassification; where values are stored pre-classified, harmonization is impossible and analyses should be reported separately per database.
3. **Search all plausible source tables before declaring a variable unavailable**. In eICU-CRD, CRT was absent from nurseCharting but present in nurseAssessment, with perfusion-related observations additionally available in physicalExam; state–site conflation within a single field requires explicit parsing rules.
4. **Treat sparsely documented populations as selected populations**. Where coverage is low, compare documented versus undocumented stays on severity markers before using the subset for model development or external validation; declare the selection explicitly when reporting.
5. **Pre-specify value-aggregation and time-window rules against empirical recording distributions**. A worst-value rule is well supported where repeated bilateral records exist, but the chosen predictor window (e.g., 0–24 h) must be checked against when records actually occur; windows that capture only a minority of records (∼29% in both databases) compound selection with sparse early ascertainment.

Beyond CRT, the five-dimensional framework used here (availability, thresholds, recording patterns and time windows, bilateral consistency, and selection bias) is transferable to other routinely documented bedside variables (for example, neurological assessments or skin findings) and to other database pairs. The absence of structured peripheral perfusion index (PPI) fields in both databases identifies a concrete target for future data collection standards [25].

### 4.5 Strengths and limitations

Strengths of this study include its scale (nearly 300,000 adult ICU stays across a single-center high-coverage database and a multi-center sparse-coverage database, a complementary structure that makes the heterogeneity visible), its reproducibility (SHA256-verified source data, documented SQL extraction logic, and a preregistered analysis program as part of the parent project), and its deliberately outcome-free scope, which allowed operational heterogeneity to be characterized without the risk of outcome-driven analytic choices.

Several limitations apply. The analysis is descriptive and no causal or predictive inferences should be drawn.

The two databases cover different calendar periods (MIMIC-IV 2008–2022; eICU-CRD 2014–2015, whose period falls within it), so secular trends in documentation practice cannot be separated from database identity. Low eICU-CRD coverage may also partly reflect heterogeneous hospital contributions, and hospital-level stratification was not performed. Both databases are US-based, and our findings may not extend to other EHR ecosystems, although the analytical framework is system-agnostic.

Standardized severity scores (e.g., APACHE IV; Sequential Organ Failure Assessment, SOFA) were not systematically available in the MIMIC-IV chartevents table, so its selection-bias analysis used hospital mortality and ICU length of stay as proxies for illness severity; APACHE IV-based comparison was possible only in eICU-CRD.

Availability rests on different windows (MIMIC-IV: first 24 hours; eICU-CRD: entire ICU stay; Table 1), a difference that is conservative for the availability gap reported here.

The stored data place limits on what can be resolved: “not measured” and “measured but not recorded” are indistinguishable [27], and the semantic equivalence of the eICU-CRD “normal” and “< 2 seconds” values cannot be confirmed from the stored data alone. Record timestamps are also subject to documentation and entry errors; extreme pre-admission offsets (for example, the MIMIC-IV record approximately one year before ICU admission) should be regarded as likely timestamp artifacts, and no plausibility filtering was applied.

## 5. Conclusions

CRT is operationalized in fundamentally different ways across two of the most widely used public ICU databases. A 73.5-percentage-point availability gap, discordant abnormality thresholds (3 vs 2 seconds), differing per-stay documentation volume (7.0 records per stay in the first 24 hours in MIMIC-IV versus 27.5 across the entire ICU stay in eICU-CRD), non-comparable bilateral structures, and opposing selection patterns (severity-driven documentation in eICU-CRD versus routine practice in MIMIC-IV). Together, these differences mean that cross-database CRT research cannot assume definitional equivalence. These conclusions derive from a descriptive analysis of two US-based databases. Studies using these databases should report availability and definitional heterogeneity explicitly, avoid pooling across thresholds, treat sparsely documented subsets as selected populations, and pre-specify aggregation rules against empirical recording patterns. Standardized common data elements for bedside perfusion assessment (including structured peripheral perfusion index fields, currently absent from both databases) are needed to make future cross-database research on CRT valid by design.

## Supporting information

STROBE_checklist

RECORD checklist

CRT item identifier and attribute mapping across databases

eICU-CRD nurseAssessment schema and CRT state/site parsing rules

Preregistration amendment note

SQL queries and Python analysis scripts

## Data Availability

All data referred to in this study are publicly available through PhysioNet (MIMIC-IV v3.1 and eICU-CRD v2.0, subject to credentialed access). SQL queries, Python analysis scripts, and the complete analytical pipeline are available online at https://doi.org/10.17605/OSF.IO/JUZNQ (Open Science Framework) and as Supplementary file 6.

https://physionet.org/content/mimiciv/3.1/

https://physionet.org/content/eicu-crd/2.0/

https://doi.org/10.17605/OSF.IO/JUZNQ

## List of abbreviations

APACHE: Acute Physiology and Chronic Health Evaluation
CI: confidence interval
CRT: capillary refill time
EHR: electronic health record
eICU-CRD: eICU Collaborative Research Database
ICU: intensive care unit
IQR: interquartile range
MIMIC-IV: Medical Information Mart for Intensive Care IV
OSF: Open Science Framework
PPI: peripheral perfusion index
RECORD: REporting of studies Conducted using Observational Routinely-collected health Data
SD: standard deviation
SMD: standardized mean difference
SOFA: Sequential Organ Failure Assessment
SQL: structured query language
STROBE: Strengthening the Reporting of Observational Studies in Epidemiology

## Declarations

### Ethics approval and consent to participate

This study is a secondary analysis of two existing, de-identified public critical care datasets, MIMIC-IV v3.1 and eICU-CRD v2.0, obtained through PhysioNet under its credentialed data use agreement (PhysioNet Credentialed Health Data Use Agreement 1.5.0). No new human-subjects data were collected, no participant was contacted and no identifiable information was accessed. The creation of MIMIC-IV was reviewed by the Institutional Review Board at the Beth Israel Deaconess Medical Center, which approved the data resource and granted a waiver of informed consent (PhysioNet, MIMIC-IV v3.1, Ethics). eICU-CRD is de-identified to the Safe Harbor standard of the US Health Insurance Portability and Accountability Act, a standard certified by Privacert(HIPAA certification no. 1031219-2) (PhysioNet, eICU-CRD v2.0, Methods). Informed consent for participation was not applicable.

### Consent for publication

Not applicable.

### Availability of data and materials

All data analyzed in this study are publicly available through PhysioNet. SQL queries, Python analysis scripts, and the complete analytical pipeline are publicly available at the Open Science Framework (https://doi.org/10.17605/OSF.IO/JUZNQ; Supplementary file 6).

### Authors’ contributions

Xia X participated in study conception and design, statistical analysis, and drafting the manuscript. Xie Z participated in study conception and design, database application and download, data cleaning and preliminary analysis, and critically revised and finalized the manuscript. All authors read and approved the final manuscript.

### Use of AI-assisted technologies in manuscript preparation

During the preparation of this manuscript, the authors used DeepSeek-V4 Pro (DeepSeek; accessed August–September 2026) to assist with language editing and with drafting SQL extraction queries and Python data-processing scripts (Section 2.8). All AI-assisted content was reviewed and verified by the authors, who take full responsibility for the integrity and accuracy of the final manuscript.

## Acknowledgements

Not applicable.

## Preregistration

This study was preregistered on the Open Science Framework (DOI: 10.17605/OSF.IO/JUZNQ) as the methodological aim (Aim 1) of the parent project, which additionally registered a prediction-modeling aim to be reported separately.

## Supplementary material

- **Supplementary file 1: STROBE checklist**. File name: Supplementary_file_1_STROBE_checklist.pdf (PDF). STROBE checklist, with the location in the manuscript of every item.
- **Supplementary file 2: RECORD checklist**. File name: Supplementary_file_2_RECORD_checklist.pdf (PDF). RECORD checklist (extension for studies conducted using routinely collected health data).
- **Supplementary file 3: CRT item identifier and attribute mapping across databases**. File name: Supplementary_file_3_CRT_itemid_cellattribute_mapping.pdf (PDF). CRT item identifier and attribute mapping across databases, including the denominator conventions underlying Tables 1–3.
- **Supplementary file 4: eICU-CRD nurseAssessment schema and CRT state/site parsing rules**. File name: Supplementary_file_4_eICU_nurseAssessment_schema.pdf (PDF). eICU-CRD nurseAssessment schema and CRT state/site parsing rules.
- **Supplementary file 5: Preregistration amendment note**. File name: Supplementary_file_5_amendment_note.pdf (PDF). Preregistration amendment note (scope adjustment and numeric discrepancies).
- **Supplementary file 6: SQL queries and Python analysis scripts**. File name: Supplementary_file_6_code.zip (ZIP). SQL queries and Python analysis scripts for all analyses (complete reproducibility package).

