## Supplementary material for "Operational Heterogeneity of Capillary Refill Time Across Two Public ICU Databases: A Methodological Analysis": STROBE_checklist

### Additional file 1: STROBE checklist

**Journal:** BMC Medical Research Methodology **Page references:** page numbers refer to the double-spaced manuscript PDF (30 pages; continuous line numbering applied).

**Study design:** Retrospective, descriptive, cross-database methodological analysis of routinely collected ICU data (two independent databases; no outcome modelling).

**STROBE reference:** von Elm E, Altman DG, Egger M, et al. The Strengthening the Reporting of Observational Studies in Epidemiology (STROBE) statement: guidelines for reporting observational studies. *Lancet*. 2007;370(9596):1453-1457. doi:10.1016/S0140-6736(07)61602-X.

> **Note on applicability.** This study characterizes how a routinely documented bedside variable (CRT) is *operationalized* in two databases. It is deliberately **outcome-free**: no exposure-outcome association is estimated, no prediction model is fitted, and no hypothesis test is performed (Methods §2.6-§2.7; Discussion §4.5, limitation 1). STROBE items that presuppose an outcome analysis are therefore recorded as "Not applicable", with the reason given, rather than left blank.

| Item no. | STROBE item | Reported in manuscript |
|---|---|---|
| **Title and abstract** | | |
| 1a | Indicate the study's design with a commonly used term in the title or the abstract | Title ("A Methodological Analysis"); Abstract, Methods sentence ("retrospective, descriptive, cross-database methodological analysis") (p. 1) |
| 1b | Provide in the abstract an informative and balanced summary of what was done and what was found | Abstract — Background, Methods, Results, Conclusions (p. 1) |
| **Introduction** | | |
| 2 | Explain the scientific background and rationale for the investigation being reported | §1 Background, paragraphs 1-3 (CRT endorsement in the 2026 Surviving Sepsis Campaign [1], ANDROMEDA-SHOCK-2 [2], prognostic evidence [3,4], and the growing use of MIMIC-IV [5] / eICU-CRD [6] for multi-database CRT research) (p. 3) |
| 3 | State specific objectives, including any prespecified hypotheses | §1 Background, final paragraph (five prespecified dimensions of operational heterogeneity); prespecification documented in the OSF registration (§2.5; Declarations, Preregistration) (pp. 3, 6, 21) |
| **Methods** | | |
| 4 | Present key elements of study design early in the paper | §2.1 Study design and setting (p. 4) |
| 5 | Describe the setting, locations, and relevant dates, including periods of recruitment, exposure, follow-up, and data collection | §2.1 (MIMIC-IV: single centre, Beth Israel Deaconess Medical Center, Boston MA, admissions 2008-2022; eICU-CRD: >200 US hospitals, 2014-2015; both extracted 2026-09-02) (p. 4) |

| Item no. | STROBE item | Reported in manuscript |
|---|---|---|
| 6a | Give the eligibility criteria, and the sources and methods of selection of participants | §2.2 Participants (all adult ICU stays; MIMIC-IV: 0 of 364,627 patients with anchor_age < 18; eICU-CRD: age >= 18 after mapping age > 89 to 90, 625 stays excluded); Figure 1 (flow diagram) (pp. 5, 27) |
| 6b | For matched studies, give matching criteria and the number of exposed and unexposed | Not applicable — no matched design; CRT-available and CRT-unavailable stays are compared descriptively without matching (§2.6) (p. 7) |
| 7 | Clearly define all outcomes, exposures, predictors, potential confounders, and effect modifiers. Give diagnostic criteria, if applicable | §2.3 (CRT definitions, both databases), §2.4 (supplementary perfusion variables); Table 1. No outcome, confounder, or effect modifier is defined because no association is estimated (pp. 5, 6, 27) |
| 8 | For each variable of interest, give sources of data and details of methods of assessment (measurement). Describe comparability of assessment methods if there is more than one group | §2.1, §2.3, §2.5; Table 1; Additional file 3 (itemid / cellattribute mapping) and Additional file 4 (eICU schema and parsing rules) (pp. 4–6, 27) |
| 9 | Describe any efforts to address potential sources of bias | §2.6 (selection bias quantification in both databases); §3.6 and Table 4 (results); §4.2 Selection bias (interpretation, including opposing directions of selection); §4.5 limitation 3 ("not measured" vs "measured but not recorded") (pp. 7, 10, 13, 19, 28) |
| 10 | Explain how the study size was arrived at | §2.2 — all eligible adult ICU stays in each database were included (MIMIC-IV 94,458; eICU-CRD 200,234); no sampling and no a priori sample-size calculation, because the analyses are descriptive and census-based rather than inferential (p. 5) |
| 11 | Explain how quantitative variables were handled in the analyses. If applicable, describe which groupings were chosen and why | §2.7 (continuous measures as mean with SD, with medians and IQR reported where distributions are skewed; Table 1); §2.3 (CRT dichotomized at each database's native threshold: 3 s in MIMIC-IV, 2 s in eICU-CRD, and the reason harmonization is impossible is given in §4.2 Thresholds) (pp. 5, 7, 13, 27) |
| 12a | Describe all statistical methods, including those used to control for confounding | §2.7 — descriptive statistics only; percent agreement and Cohen kappa for bilateral consistency; standardized mean differences for group comparisons. No confounder control (no effect estimation) (p. 7) |
| 12b | Describe any methods used to examine subgroups and interactions | Not applicable — no subgroup or interaction analysis was performed |
| 12c | Explain how missing data were addressed | §2.2, §3.2 — no imputation was performed. Absence of CRT records is the object of study, not a nuisance to be completed; CRT-available and CRT-unavailable stays are compared explicitly (§2.6, §3.6). Within-record missingness is reported through the availability denominators in Tables 1-3 (pp. 5, 7, 8, 10, 27) |
| 12d | If applicable, explain how loss to follow-up was addressed | Not applicable — no longitudinal follow-up; each ICU stay is characterized independently |

| Item no. | STROBE item | Reported in manuscript |
|---|---|---|
| 12e | Describe any sensitivity analyses | Not applicable — no modelling was performed; the only analytic choice relevant to results (value-aggregation rule) is discussed in §4.2 Recording patterns and §4.4 recommendation 5 (pp. 13, 18) |
| **Results** | | |
| 13a | Report the numbers of individuals at each stage of the study | §3.1, §3.2; Figure 1 (MIMIC-IV: 364,627 patients / 94,458 adult ICU stays / 75,421 CRT-available; eICU-CRD: 200,859 stays / 625 excluded / 200,234 adult stays / 12,707 CRT-available) (pp. 8, 27) |
| 13b | Give reasons for non-participation at each stage | Figure 1 and §2.2 — the single exclusion step in each database is age < 18 years (pp. 5, 27) |
| 13c | Consider use of a flow diagram | Figure 1 (ICU stay flow diagram for both databases) (p. 27) |
| 14a | Give characteristics of study participants and information on exposures and potential confounders | §3.1 (database-level composition), §3.3 (value distributions), Table 4 (age, sex, APACHE IV, acute physiology score, predicted ICU mortality, actual hospital mortality by CRT availability); MIMIC-IV comparison in the Table 4 note (age, sex, hospital mortality, ICU length of stay) (pp. 8, 9, 28) |
| 14b | Indicate the number of participants with missing data for each variable of interest | §3.2 and Table 1 — CRT availability is reported for every stay (79.85% vs 6.35%), which is the primary completeness measure; supplemental perfusion variable coverage is given in Table 5 (pp. 8, 27, 29) |
| 14c | Summarize follow-up time | Not applicable — no follow-up period. ICU length of stay is reported descriptively in §3.6 and the Table 4 note (pp. 10, 28) |
| 15 | Report numbers of outcome events or summary measures | Not applicable — the study is outcome-free by design (§4.5, limitation 1; §4.4). Outcome-free scope is a stated methodological feature, not an omission (pp. 18, 19) |
| 16 | Give unadjusted estimates and, if applicable, confounder-adjusted estimates and their precision; report category boundaries for categorized variables | §3.2-§3.8 and Tables 1-5 — all reported quantities are descriptive estimates (proportions, means with SD, medians with IQR, Cohen kappa, SMD). No adjusted estimates or precision intervals exist because no effect is estimated. Category boundaries are the native thresholds of each database and are stated explicitly (§2.3, §4.2 Thresholds) (pp. 5, 8, 12, 13, 27) |
| 17 | Report other analyses done | §3.6 (selection bias in both databases), §3.7 (bilateral consistency), §3.8 (hands/feet site concordance and supplementary perfusion variables), §3.9 (cross-database definitional mapping) (pp. 10–12) |
| **Discussion** | | |
| 18 | Summarise key results with reference to study objectives | §4.1 Summary of principal findings (p. 13) |

| Item no. | STROBE item | Reported in manuscript |
|---|---|---|
| 19 | Discuss limitations, taking into account sources of potential bias or imprecision. Discuss both direction and magnitude of any potential bias | §4.5 Strengths and limitations (eight numbered limitations, including calendar-period confounding, indistinguishable "not measured" vs "not recorded", proxy severity measures in MIMIC-IV, unresolved semantic equivalence of eICU-CRD state labels, temporal misalignment, non-stratified hospital composition, and US-only provenance); direction and magnitude of selection are quantified in §3.6 and §4.2 (pp. 10, 13, 19) |
| 20 | Give a cautious overall interpretation of results considering objectives, limitations, multiplicity of analyses, results from similar studies, and other relevant evidence | §4.2 (interpretation by dimension), §4.3 (comparison with previous work, including the distinction between documentation consistency and measurement reliability) (pp. 13, 16) |
| 21 | Discuss the generalisability (external validity) of the study results | §4.4 (transferability of the five-dimensional framework to other bedside variables and database pairs), §4.5 limitation 8 (US-based databases; framework is system-agnostic) (pp. 18, 19) |
| **Other information** | | |
| 22 | Give the source of funding and the role of the funders | Declarations — Funding (no specific grant; funders had no role); see also Additional file 2 (RECORD 22.1) (p. 21) |

**Reference for this checklist:** STROBE Statement — checklist of items that should be addressed in reports of observational studies (cohort, case-control, and cross-sectional studies combined). Available at https://www.strobe-statement.org/checklists/ (accessed 2026-09-10).

**Companion file:** Additional file 2 — RECORD checklist (extension for studies using routinely collected health data).
