## Supplementary material for "Operational Heterogeneity of Capillary Refill Time Across Two Public ICU Databases: A Methodological Analysis": RECORD checklist

### Additional file 2: RECORD checklist

**Journal:** BMC Medical Research Methodology **Page references:** page numbers refer to the double-spaced manuscript PDF (30 pages; continuous line numbering applied).

**RECORD reference:** Benchimol EI, Smeeth L, Guttmann A, et al. The REporting of studies Conducted using Observational Routinely-collected health Data (RECORD) statement. *PLoS Med*. 2015;12(10):e1001885. doi:10.1371/journal.pmed.1001885.

> **Note on applicability.** RECORD extends STROBE for studies using routinely collected health data. Items referring to cross-database **linkage** are not applicable: MIMIC-IV and eICU-CRD were analysed independently and no person-level linkage was performed or attempted (the databases contain no shared patient identifiers). This is stated here so that the "no linkage" status is explicit rather than implicit.

| RECORD item | Item text (RECORD statement) | Reported in manuscript / supplement |
|---|---|---|
| **Title and abstract** | | |
| RECORD 1.1 | The type of data used should be specified in the title or abstract. When possible, the name of the databases used should be included | Title ("Across Two Public ICU Databases"); Abstract, Methods ("MIMIC-IV v3.1 ... and eICU-CRD v2.0") (p. 1) |
| RECORD 1.2 | If applicable, the geographic region and time frame within which the study took place should be reported in the title or abstract | Abstract, Methods and Results (MIMIC-IV: single-centre US data, 2008-2022; eICU-CRD: >200 US hospitals, 2014-2015); §2.1 (pp. 1, 4) |
| RECORD 1.3 | If linkage between databases was conducted for the study, this should be clearly stated in the title or abstract | Not applicable — no linkage between databases was performed |
| **Introduction** | | |
| RECORD 2.1 | Explain the scientific background and rationale for using the study design and the data sources | §1 Background, paragraphs 2-3 (CRT endorsement and trial evidence, and the specific rationale for using MIMIC-IV and eICU-CRD: complementary single-centre high-coverage vs multi-centre sparse-coverage structures) (p. 3) |
| RECORD 3.1 | State the specific objectives, including whether the objectives were prespecified or exploratory; if a protocol exists, state where it can be accessed | §2.5 (registration statement), Declarations — Preregistration, and Additional file 5 (amendment note). Objectives were prespecified: the methodological aim (Aim 1) of the parent project, registered on OSF before the analyses reported here (DOI: 10.17605/OSF.IO/JUZNQ). A second registered aim (prediction modelling) is deferred and reported separately (pp. 6, 21) |
| **Methods** | | |
| RECORD 4.1 | The study design, including the type of data used, should be stated | §2.1 (retrospective, descriptive, cross-database methodological analysis of routinely collected EHR data) (p. 4) |

| RECORD item | Item text (RECORD statement) | Reported in manuscript / supplement |
|---|---|---|
| RECORD 5.1 | The settings and locations where the data were collected, and eligibility criteria, should be reported | §2.1 (settings and locations), §2.2 (eligibility: adult ICU stays; no treatment or time-window restrictions) (pp. 4, 5) |
| RECORD 6.1 | The methods of study population selection (such as codes or algorithms used to identify subjects) should be listed in detail. If this is not possible, an explanation should be provided | §2.2 (adult-stay definitions, including mapping of eICU-CRD age > 89 to 90); §2.3.1 and §2.3.2 (CRT identification algorithms); Additional files 3 and 4; code in `code/sql/01_extract_mimic.sql` and `code/sql/02_extract_eicu.sql` (p. 5) |
| RECORD 6.2 | Any validation studies of the codes or algorithms used to select the population should be referenced. If validation was conducted for this study and not published elsewhere, detailed methods and results should be provided | §2.3.1 (systematic search of the MIMIC-IV `d_items` dictionary; no further itemids identified), §3.3 (observed value distribution of every retrieved category), §2.8 (AI-assisted code validated against source record counts and by manual review of SQL logic). No external criterion standard exists for either database's CRT field, so validation was limited to internal value-structure verification; this is stated as a limitation in §4.5 (limitation 5) (pp. 5, 8, 9, 19) |
| RECORD 6.3 | If the study involved linkage of databases, consider use of a flow diagram or other graphical display to demonstrate the data linkage process, including the number of individuals with linked data at each stage | Not applicable — no linkage. A study flow diagram is nevertheless provided (Figure 1), showing numbers at each selection stage in both databases (p. 27) |
| RECORD 7.1 | A complete list of codes and algorithms used to classify exposures, outcomes, confounders, and effect modifiers should be provided. If these cannot be reported, an explanation should be provided | Additional file 3 (MIMIC-IV itemids 223951/224308; eICU-CRD `cellattribute` "Capillary Refill" with its full value taxonomy; threshold mapping; aggregation rules) and Additional file 4 (eICU-CRD state/site parsing algorithm); SQL in `code/sql/` |
| RECORD 12.1 | Authors should describe the extent to which the investigators had access to the database population used to create the study population | §2.1 (both databases accessed through PhysioNet under the required data use agreements; full local copies of the population tables); Declarations — Availability of data and materials; `code/README.md` (data access section) (pp. 4, 21) |
| RECORD 12.2 | Authors should provide information on the data cleaning methods used in the study | §2.1 (SHA256 verification of all source files after download), §2.2 (age > 89 mapping; adult filtering), §2.3.2 and Additional file 4 (state/site parsing rules and exclusions), §2.5 (timestamp conversion and classification of pre-admission records), §2.8 (verification of code outputs against source record counts); `code/README.md` (pp. 4–6, 8) |
| RECORD 12.3 | State whether the study included person-level, institutional-level, or other data linkage across two or more databases. The methods of linkage and methods of linkage quality evaluation should be provided | Not applicable — no linkage. MIMIC-IV and eICU-CRD were analysed separately; all cross-database statements are comparisons of independently derived summary measures. Database composition differs at the hospital level (single centre vs >200 hospitals) and this is discussed in §4.5 (limitation 7) (p. 19) |
| **Results** | | |

| RECORD item | Item text (RECORD statement) | Reported in manuscript / supplement |
|---|---|---|
| RECORD 13.1 | Describe in detail the selection of the persons included in the study, including filtering based on data quality, data availability, and linkage. The selection of included persons can be described in the text and/or by means of the study flow diagram | §3.1, §3.2 and Figure 1 — from source populations to adult ICU stays to CRT-available stays in each database, including the availability ("data availability") step that defines the analysed subgroups (pp. 8, 27) |
| **Discussion** | | |
| RECORD 19.1 | Discuss the implications of using data that were not created or collected to answer the specific research question(s). Include discussion of misclassification bias, unmeasured confounding, missing data, and changing eligibility over time as they pertain to the study being reported | §4.5 (limitation 3: "not measured" and "measured but not recorded" are indistinguishable; limitation 4: proxy severity measures in MIMIC-IV; limitation 6: differing recording windows; limitation 7: hospital-level composition) and §4.2 (Selection bias; misclassification under discordant thresholds; and the explicit distinction between documented CRT and protocolized CRT) (pp. 13, 19) |
| **Other information** | | |
| RECORD 22.1 | Authors should provide information on how to access any supplemental information such as the study protocol, raw data, or programming code | Declarations — Availability of data and materials (OSF repository, DOI: 10.17605/OSF.IO/JUZNQ) and Preregistration; Additional files 3-5 and the code bundle in `code/`; source data are available from PhysioNet under the respective data use agreements and are not redistributable (p. 21) |

**Reference for this checklist:** RECORD statement — checklist of items, extended from the STROBE statement, that should be reported in observational studies using routinely collected health data (Benchimol et al., *PLoS Med* 2015;12(10):e1001885). Checklist text reproduced under CC BY 4.0.

**Companion file:** Additional file 1 — STROBE checklist.
