## Supplementary material for "Operational Heterogeneity of Capillary Refill Time Across Two Public ICU Databases: A Methodological Analysis": CRT item identifier and attribute mapping across databases

### Additional file 3: CRT item identifier and attribute mapping across databases

**Purpose.** This file documents, in reproducible detail, how capillary refill time (CRT) was located and mapped in each database: the dictionaries searched, the search terms, the identifiers and attribute values retrieved, the binary threshold applied, the value and stay distributions actually observed, and the denominator conventions used in each table of the manuscript. It supports Methods §2.3 and Results §3.3, §3.4 and §3.9, and reporting items STROBE 7-8 and RECORD 6.1, 6.2, 7.1.

#### 1. MIMIC-IV v3.1

##### 1.1 Variable location procedure

| Step | Action | Result |
|---|---|---|
| 1 | Systematic search of the `d_items` dictionary (ICU module) for labels containing `capillary` or `refill` | Two itemids retrieved — no other candidate item was found |
| 2 | Verification of the retrieved itemids in `chartevents` | Both populated; values are categorical strings |
| 3 | Verification of remaining candidates | None. No additional itemid in `d_items` matched the search terms |

> Reported in §2.3.1: "CRT was identified through a systematic search of the `d_items` dictionary for labels containing 'capillary' or 'refill,' which yielded two itemids: 223951 (right hand) and 224308 (left hand) in the `chartevents` table. No additional itemids were identified."

##### 1.2 Identifier and value mapping

| Element | Value |
|---|---|
| Source table | `chartevents` (ICU module) |
| Dictionary | `d_items` (ICU module) |
| Search terms | `capillary`, `refill` (case-insensitive, substring match on `label`) |
| Itemid — right hand | `223951` (Capillary Refill Time [Right Hand]) |
| Itemid — left hand | `224308` (Capillary Refill Time [Left Hand]) |
| Stored value — normal | `Normal <3 Seconds` |
| Stored value — abnormal | `Abnormal >3 Seconds` |

| Element | Value |
| --- | --- |
| Value type | Pre-classified binary string (no numeric value stored) |
| Operative threshold | 3 seconds |
| Sides | Separate itemids for left and right hand; no feet equivalent |

##### 1.3 Threshold boundary note

The stored labels encode strict inequalities (`<3` and `>3`). A measurement of exactly 3 seconds is not assigned to either stored category by the source encoding, and no record was found in an unclassifiable state. This boundary is therefore undefined in the source data, not resolved by the analyst; because no numeric CRT value is stored, no alternative boundary can be applied. Stated for completeness.

##### 1.4 Observed distribution (MIMIC-IV)

| Quantity | Value | Denominator / window |
| --- | --- | --- |
| Adult ICU stays | 94,458 | all adult ICU stays |
| CRT-available stays | 75,421 (79.85%) | ≥ 1 CRT record within the first 24 h of ICU admission |
| CRT records, all course, both sides | 1,892,151 | all records across all stays |
| CRT records within the first 24 h | 542,782 (28.7% of 1,892,151) | includes pre-admission records (0.7%) |
| Abnormal records | 3.7% of records | all records, both sides |
| Mean records per stay | 7.0 (median 6, IQR 4-10) | first 24 h, 75,421 CRT-available stays |
| Bilateral record availability | 99.6% | 75,421 CRT-available stays |
| Left-right agreement | 99.4% (Cohen's kappa = 0.914) | bilateral stays, most recent record per side |
| Discordant bilateral stays | 430 (0.6%) | 238 right-normal/left-abnormal; 192 right-abnormal/left-normal |
| Same-timestamp bilateral pairs | 98.2% (mean absolute difference 0.12 h) | bilateral stays |

Time-window distribution by record (Table 3): pre-admission 12,871 (0.7%); 0-6 h 176,199 (9.3%); 6-12 h 128,512 (6.8%); 12-24 h 225,200 (11.9%); >24 h 1,349,369 (71.3%).

##### 1.5 Aggregation and pairing rules

- **Bilateral pairing:** left- and right-hand records were paired at stay level using the most recent record of each side; pairing required at least one record per side within the first 24 h (§2.7).

- **Value aggregation for the bilateral analysis:** the binary classification of each side (normal/abnormal) at the paired time point; agreement was defined as equal binary classification on both sides.
- **Worst-value rule (deferred analysis, not applied here):** repeated measurements per stay support a worst-value summary rule for future prediction analyses; this study reports recording patterns rather than deriving a predictor (§3.4, §4.4 recommendation 5).

---

#### 2. eICU-CRD v2.0

##### 2.1 Variable location procedure

| Step | Action | Result |
|---|---|---|
| 1 | Search of `nurseCharting` for CRT content | **Zero records.** The variable is absent from this table |
| 2 | Search of `nurseAssessment` for CRT content | CRT present — retrieved via `celllabel` matching `%apillary%` |
| 3 | Verification of the attribute label | `cellattribute` = "Capillary Refill" |
| 4 | Inspection of all distinct `cellattributevalue` entries for that attribute | Mixed **state** values and **site** values in a single column (see §2.2) |

> The failed `nurseCharting` search is reported in §2.3.2 and converted into a generalizable recommendation (search all plausible source tables before declaring a variable unavailable; §4.4 recommendation 3).

##### 2.2 Value taxonomy (single attribute, two semantic classes)

| Class | `cellattributevalue` | Meaning | Records | % of 349,725 | Stays |
|---|---|---|---|---|---|
| State | `normal` | normal refill | 180,145 | 51.5% | 10,914 |
| State | `< 2 seconds` | refill < 2 s | 46,975 | 13.4% | 4,191 |
| State | `> 2 seconds` | refill > 2 s | 44,537 | 12.7% | 3,168 |
| Site | `hands` | upper-extremity site | 37,009 | 10.6% | 2,799 |
| Site | `feet` | lower-extremity site | 41,059 | 11.7% | 2,845 |
| — | **Total** | — | **349,725** | 100% | **12,707** (any CRT record) |

- **State records:** 271,657 across 12,707 stays. **Site records:** 78,068 across 3,489 stays.

- **Stay counts by category are not mutually exclusive** — a stay contributes to multiple categories across time; 12,707 is therefore less than the sum of category-specific stay counts.
- **Site co-occurrence:** of 3,489 stays with any site record, 2,155 had both hands and feet, 644 hands only, and 690 feet only (2,155 + 644 + 690 = 3,489).
- **Threshold boundary:** `normal` and `< 2 seconds` coexist as distinct stored values; their semantic equivalence cannot be established from the stored data (§3.3, §4.5 limitation 5).

##### 2.3 Binary threshold mapping applied

| Stored value | Mapped to |
|---|---|
| `normal` | normal |
| `< 2 seconds` | normal |
| `> 2 seconds` | abnormal |
| `hands`, `feet` | site attribute — no state value; contributes no binary state |

Operative threshold: **2 seconds** (conceptual; embedded in the value labels rather than stored as a number). The reason the two databases' classifications cannot be harmonized is given in §4.2 (Thresholds): both store pre-classified text, so neither can be re-derived against the other's cutoff.

##### 2.4 Observed distribution (eICU-CRD)

| Quantity | Value | Denominator / window |
|---|---|---|
| Adult ICU stays | 200,234 | after excluding 625 stays not meeting the adult criterion (age < 18 or unrecorded, from 200,859) |
| CRT-available stays | 12,707 (6.35%) | ≥ 1 CRT record, all course |
| CRT records, total | 349,725 | all course, state + site |
| Mean records per stay | 27.5 (median 13, IQR 6-29) | all course, 12,707 CRT-available stays, state + site rows |
| Records within the first 24 h | 79,165 (29.1% of 271,657 state records) | includes pre-admission records (1.1%) |
| Hands/feet concordance | 1,906 of 2,147 valid pairs (88.8%) | both sites recorded with a state value at each site |
| Discordant pairs | 241 (11.2%) | hands normal/feet abnormal 198 (9.2%); reverse 43 (2.0%) |

Time-window distribution by **state** record (Table 3): pre-admission 3,076 (1.1%); 0-6 h 22,502 (8.3%); 6-12 h 19,092 (7.0%); 12-24 h 34,495 (12.7%); >24 h 192,492 (70.9%).

##### 2.5 Aggregation and pairing rules

- **Site (hands/feet) concordance:** state and site are recorded as separate rows under the same time offset (`nurseassessoffset`). A state value recorded at an offset applies to the site(s) recorded at that offset. The hands state and feet state of a stay were taken as the most recent offset at which each site had an accompanying state value, then compared (concordant vs discordant). See Additional file 4 for the full algorithm.
- **Left/right bilateral consistency was not computed for eICU-CRD**, because its site attribute is anatomical (hands/feet) rather than lateral; the resulting quantity answers a different physiological question and has no MIMIC-IV counterpart (§3.7, §4.2 Bilateral consistency).

---

#### 3. Cross-database mapping summary

| Domain | MIMIC-IV v3.1 | eICU-CRD v2.0 | Comparable? |
|---|---|---|---|
| Source table | `chartevents` | `nurseAssessment` | No — different data structures |
| Identifier | itemid 223951 (R), 224308 (L) | `cellattribute` "Capillary Refill" | No |
| Threshold | 3 s | 2 s | No — discordant cutoffs |
| Value format | Binary (Normal/Abnormal) | Multi-state (normal, <2 s, >2 s) + site (hands/feet) | No |
| Laterality / site | left/right hand | hands/feet | No — lateral vs anatomical |
| Availability | 79.85% (first 24 h) | 6.35% (all course) | Windows differ; rates not directly comparable |
| Bilateral axis | left vs right (99.6% bilateral; 99.4% agreement, kappa 0.914) | hands vs feet (2,155 stays both sites; 88.8% concordance) | No — different axes |
| PPI (peripheral perfusion index) | Not structured | Not structured | Absent in both |

---

#### 4. Denominator conventions used in the manuscript tables

Documented here so that every count and mean in Tables 1-3 can be reproduced exactly and so that no across-table comparison is made between non-equivalent denominators.

| Table | What is counted | MIMIC-IV | eICU-CRD |
|---|---|---|---|
| Table 1 (availability) | Stays with ≥ 1 CRT record | 75,421 / 94,458 (79.85%), **first-24-h window** | 12,707 / 200,234 (6.35%), **all course** |
| Table 1 (mean records/stay) | Records per CRT-available stay | 7.0, **first 24 h** | 27.5, **all course, state + site rows** |

| Table | What is counted | MIMIC-IV | eICU-CRD |
|---|---|---|---|
| Table 2 | Recording patterns among CRT-available stays | 7.0 records/stay, 99.6% bilateral, 99.4% agreement, denominator 75,421 stays | Not applicable |
| Table 3 | **Record-level** counts by time window | All-course records, both sides: 1,892,151; within 24 h 542,782 (28.7%) | **State records only**: 271,657; within 24 h 79,165 (29.1%). Site records (78,068) carry no state value and are excluded from this table |

**Consequence for interpretation.** The mean records per stay implied by Table 3 is not comparable with the means in Table 1 or Table 2, because the windows differ (all course vs first 24 h) and, for eICU-CRD, the record types differ (state only vs state + site):

- MIMIC-IV, all course, both sides: 1,892,151 / 75,421 = **25.1** records per CRT-available stay.
- eICU-CRD, all course, state records only: 271,657 / 12,707 = **21.4** records per stay.
- eICU-CRD, all course, state + site: 349,725 / 12,707 = **27.5**, matching the 27.5 reported in Table 1.
- MIMIC-IV, first 24 h: the Table 3 window counts / 75,421 give **7.2**; the Table 1 and Table 2 mean is **7.0**. Both refer to the first-24-h window and differ only in how window-edge and repeated-timestamp records are handled between the per-stay analysis dataset (Table 1/2) and the record-level window tabulation (Table 3).

The availability percentages for the two databases in Table 1 are likewise **not** computed over the same window (first 24 h vs all course), and this is stated in the Table 1 footnote, §3.2 and §4.5 limitation 9.

---

#### 5. Provenance

| Artefact | Location |
|---|---|
| MIMIC-IV extraction SQL | `code/sql/01_extract_mimic.sql` |
| eICU-CRD extraction SQL | `code/sql/02_extract_eicu.sql` |
| Availability analysis | `code/scripts/03_availability.py` |
| Recording-pattern analysis | `code/scripts/04_recording_patterns.py` |
| Bilateral consistency (MIMIC-IV) | `code/scripts/05_bilateral_mimic.py` |
| Site concordance (eICU-CRD) | `code/scripts/06_site_concordance_eicu.py` |
| Selection bias (eICU-CRD / MIMIC-IV) | `code/scripts/07_selection_bias_eicu.py`, `code/scripts/08_selection_bias_mimic.py` |
| Pipeline entry point | `code/run_all.py` |
| Public repository | https://doi.org/10.17605/OSF.IO/JUZNQ |
