## Supplementary material for "Operational Heterogeneity of Capillary Refill Time Across Two Public ICU Databases: A Methodological Analysis": eICU-CRD nurseAssessment schema and CRT state/site parsing rules

### Additional file 4: eICU-CRD nurseAssessment schema and CRT state/site parsing rules

**Purpose.** The eICU-CRD `nurseAssessment` table stores CRT as one attribute whose values mix a **clinical state** and an **anatomical site** in a single column. Recovering "hands vs feet" or "normal vs abnormal" therefore requires an explicit parsing step that a naive `GROUP BY cellattributevalue` would get wrong. This file formalizes the table structure actually used, the parsing rules, and the pairing algorithm behind Results §3.3, §3.8 and the hands/feet concordance figures. It supports STROBE item 8 and RECORD items 6.1, 7.1 and 12.2.

---

#### 1. Table structure as used

| Column | Role in this analysis |
|---|---|
| `patientunitstayid` | ICU stay identifier (unit of analysis) |
| `nurseassessoffset` | Minutes from ICU admission — the time axis used for windows and for pairing |
| `cellattribute` | Attribute name; the CRT attribute is **"Capillary Refill"** |
| `cellattributevalue` | The stored value — **mixes state and site semantics** (see §2) |
| `celllabel` | Display label; CRT rows were retrieved by `celllabel LIKE '%apillary%'` |
| `cellattributepath` | Hierarchical parent path for the attribute, as documented in the eICU-CRD v2.0 data dictionary; not used as a filter in this analysis |

**Retrieval predicate actually applied:** `WHERE celllabel LIKE '%apillary%'` (yielding the "Capillary Refill" attribute). The corresponding `cellattribute` value was verified to be "Capillary Refill".

**Negative result worth recording:** the same content is **not** present in `nurseCharting` — a search of that table returned zero CRT records. Studies that search only `nurseCharting` will conclude, incorrectly, that CRT does not exist in eICU-CRD (§2.3.2, §4.4 recommendation 3).

---

#### 2. The state/site mixing problem

For a given stay and `nurseassessoffset`, CRT is not stored as one row with a state and a site. It is stored as **independent rows** in the same attribute:

```
patientunitstayid | nurseassessoffset | cellattribute     | cellattributevalue
------------------+-------------------+-------------------+-------------------
123456            | 1440              | Capillary Refill  | normal
123456            | 1440              | Capillary Refill  | hands
123456            | 1440              | Capillary Refill  | feet
```

Consequences:

1. **A state value recorded at an offset applies to the site(s) recorded at that offset** — the two rows are concurrent, not alternative values of one variable.
2. `cellattributevalue` cannot be grouped as a single categorical variable: doing so counts "hands" as if it were a third clinical state alongside "normal" and "> 2 seconds".
3. Stay-level counts by value are **not mutually exclusive** (a stay contributes to as many categories as it has across time).
4. Only rows carrying a state value carry clinical information; site rows alone contribute location, not severity.

| **Semantic class** | **`cellattributevalue` values** | **Count in the analysed extract** |
| --- | --- | --- |
| State | `normal`, `< 2 seconds`, `> 2 seconds` | 271,657 records |
| Site | `hands`, `feet` | 78,068 records |
| — | Total CRT-related rows | 349,725 records |

With threshold mapping: `normal` and `< 2 seconds` → normal; `> 2 seconds` → abnormal (§2.3.2, §4.2 Thresholds).

---

#### 3. Parsing pseudocode

```
# Step 1 - retrieve the attribute
crt_all = SELECT patientunitstayid, nurseassessoffset, cellattributevalue
          FROM   nurseAssessment
          WHERE  celllabel LIKE '%apillary%'

# Step 2 - classify each row as state or site (semantic parsing, not a lookup of IDs)
for each row:
    if value in ('normal', '< 2 seconds')  -> binary_state = 'normal',   site = NULL
    if value == '> 2 seconds'              -> binary_state = 'abnormal', site = NULL
    if value == 'hands'                    -> site = 'hands',  binary_state = NULL
    if value == 'feet'                     -> site = 'feet',   binary_state = NULL

# Step 3 - collapse to one state per (stay, offset) and flag which sites are present
offset_states = GROUP BY (patientunitstayid, nurseassessoffset)
                SELECT MAX(binary_state)     AS state_value   WHERE binary_state IS NOT NULL
offset_sites  = GROUP BY (patientunitstayid, nurseassessoffset)
                SELECT MAX(site='hands')     AS has_hands,
                       MAX(site='feet')      AS has_feet     WHERE site IS NOT NULL

# Step 4 - restrict to stays with both sites recorded
stays_both = stays having COUNT(DISTINCT site) = 2      # 2,155 stays

# Step 5 - join state to site at the same offset, then take the most recent
#          offset at which each site has an accompanying state value
site_states  = offset_states JOIN offset_sites USING (patientunitstayid, nurseassessoffset)
hands_latest = most recent (ORDER BY nurseassessoffset DESC) site_states row with has_hands
feet_latest  = most recent row with has_feet
paired       = hands_latest JOIN feet_latest USING (patientunitstayid)   # 2,147 valid pairs

# Step 6 - classify each pair
concordant  : state(hands) == state(feet)                                # 1,906 (88.8%)
discordant  : state(hands) != state(feet)                                # 241   (11.2%)
```

**Why the most-recent rule:** state and site are not recorded at every offset for every site, so requiring an exact-offset match between a specific hands row and a specific feet row would discard most stays. Taking the most recent offset at which each site has a concurrent state value keeps the comparison within the limit of what the data support. The price is that hands and feet states may come from different offsets; the resulting quantity is therefore a **documentation** concordance and not a simultaneous two-site measurement.

#### 4. Worked reconciliation of the reported numbers

| Step | Stays | Records | Source |
|---|---|---|---|
| Any CRT row (state or site) | 12,707 | 349,725 | Table 1 |
| Rows carrying a state value | 12,707 | 271,657 | Table 3 denominator |
| Rows carrying a site value | 3,489 | 78,068 | §3.3 |
| Both hands and feet recorded | 2,155 | — | §3.3 |
| Hands only | 644 | — | §3.3 |
| Feet only | 690 | — | §3.3 |
| Both-sites stays with a valid state at each site | 2,147 | — | §3.8 |
| Concordant pairs | 1,906 (88.8%) | — | §3.8 |
| Discordant pairs | 241 (11.2%) | — | §3.8 |
| Discordant: hands normal / feet abnormal | 198 (9.2%) | — | §3.8 |
| Discordant: hands abnormal / feet normal | 43 (2.0%) | — | §3.8 |

Checks: 2,155 + 644 + 690 = 3,489 stays with any site record; 271,657 + 78,068 = 349,725 rows; 1,906 + 241 = 2,147 valid pairs; 198 + 43 = 241 discordant. The directional asymmetry (198 vs 43) — distal lower-extremity recordings abnormal while hands are normal — is discussed in §3.8 and §4.2.

---

#### 5. Timing and window handling

- All windows are computed from `nurseassessoffset` (minutes from ICU admission), converted to hours.
- Negative offsets (before ICU admission) are classified as **pre-admission** rather than discarded: 1.1% of eICU-CRD state records.
- The eICU-CRD stay-level admission field carries limited precision, which is why stay-level availability in eICU-CRD is reported as all-course rather than first-24-h, while MIMIC-IV availability is first-24-h. This asymmetry is stated in the Table 1 footnote, §3.2 and §4.5 limitation 9.

---

#### 6. Provenance

| Artefact | Location |
|---|---|
| Extraction SQL (state/site parsing) | `code/sql/02_extract_eicu.sql` |

| Artefact | Location |
|---|---|
| Site concordance algorithm | `code/scripts/06_site_concordance_eicu.py` |
| Availability and recording patterns | `code/scripts/03_availability.py`, `code/scripts/04_recording_patterns.py` |
| Selection bias | `code/scripts/07_selection_bias_eicu.py` |
| Public repository | https://doi.org/10.17605/OSF.IO/JUZNQ |
