## Supplementary material for "Operational Heterogeneity of Capillary Refill Time Across Two Public ICU Databases: A Methodological Analysis": Preregistration amendment note

### Additional file 5: Preregistration amendment note

**Date:** September 2026 (revised)

**Registered title:** Predictive Value of Capillary Refill Time for 28-Day Mortality in Septic Shock: A Cross-Database Study (MIMIC-IV and eICU-CRD)

**Submitted manuscript title:** Operational Heterogeneity of Capillary Refill Time Across Two Public ICU Databases: A Methodological Analysis

**DOI:** 10.17605/OSF.IO/JUZNQ

#### 1. Summary of scope adjustment

The preregistered parent project contained two aims:

- **Aim 1 (Methodological):** Characterize operational heterogeneity of CRT across MIMIC-IV and eICU-CRD — availability, thresholds, recording patterns, bilateral consistency, and selection bias.
- **Aim 2 (Prediction modeling):** Evaluate incremental predictive value of CRT for 28-day mortality using ΔAUC, NRI, IDI, DCA, and calibration.

**This manuscript reports Aim 1 in full as a standalone paper.** Aim 2 is deferred to a separate report.

#### 2. Rationale for separation

During execution of the analysis pipeline, the depth and methodological significance of the cross-database heterogeneity findings (73.5pp availability gap, discordant thresholds, opposing selection patterns) exceeded the scope of a supplementary methods section. Separating Aim 1 into a dedicated methodological paper provides the field with a properly validated reference on CRT data quality before prediction models relying on these data are published.

This is consistent with the parent project's registered change policy (§15), which states that any post-registration adjustments to scope will be disclosed.

#### 3. Relationship between registration and manuscript

| Element | Registration | This manuscript |
|---|---|---|
| Scope | Aim 1 only | Aim 1 only |
| Databases | MIMIC-IV v3.1 + eICU-CRD v2.0 | Same |
| Analyses | Descriptive, outcome-free | Same |
| Key findings | Feasibility probe | Full characterization with quantitative results |
| Reporting guidelines | STROBE + RECORD | Same |

All analyses in this manuscript are descriptive and methodological in nature, consistent with the feasibility-probe scope declared at registration. No causal or predictive inferences are drawn.

#### 4. Numeric discrepancies

Preliminary probe numbers in the registration documents differ from final estimates due to refined inclusion criteria, updated data processing, and corrected window definitions. Per the SAP §4.4 ("final numbers at time of analysis freeze supersede registration estimates"), the numbers in this manuscript are authoritative.

| Metric | Registration probe | Manuscript (final) | Explanation |
|---|---|---|---|
| MIMIC records/stay | 7.5 | 7.0 | Refined to first-24h window |

| Metric | Registration probe | Manuscript (final) | Explanation |
|---|---|---|---|
| MIMIC bilateral | 99.8% | 99.6% | Updated denominator |
| MIMIC agreement | 98.4% | 99.4% | Recalculated with κ |

#### 5. Prediction modeling aim (Aim 2)

Deferred to a separate report. Not included in this submission. The preregistered analysis plan for Aim 2 remains valid and will be executed as originally planned.

#### 6. Changes to registration documents

- **SAP landmark exclusion:** Corrected from −1,544 to −1,545 (21,189 − 19,644 = 1,545). The SAP contained a transcription error; the PDF version had the correct number.
- **No other changes** to the registered analysis plan.

#### 7. Data and code availability

All data analyzed in this study are publicly available through PhysioNet (MIMIC-IV v3.1 and eICU-CRD v2.0, requiring credentialing). SQL queries, Python analysis scripts, and the complete analytical pipeline are publicly available in this OSF repository, together with the preregistration and statistical analysis plan of the parent project. Final numeric estimates reported in the manuscript supersede any preliminary values in the registration documents.
