## Supplementary material for "Operational Heterogeneity of Capillary Refill Time Across Two Public ICU Databases: A Methodological Analysis": SQL queries and Python analysis scripts: README.md

# Reproducibility Package: CRT Cross-Database Methodological Analysis

**Paper:** *Operational Heterogeneity of Capillary Refill Time Across Two Public ICU Databases: A Methodological Analysis*

**Journal:** BMC Medical Research Methodology

## Quick Start

```bash
# 1. Install dependencies
pip install -r requirements.txt

# 2. Download data (see Data Access below)

# 3. Run the complete pipeline
python run_all.py --mimic-db data/processed/mimiciv_3_1.duckdb \
                  --eicu-dir /path/to/eicu_crd_2_0 \
                  --output-dir output/

# 4. Verify checksums
python verify_checksums.py --check
```

## Data Access

> **Important:** MIMIC-IV and eICU-CRD are governed by PhysioNet Data Use Agreements (DUA) and **must not be redistributed**. This reproducibility package contains only analysis code — raw data files are not included.

- **MIMIC-IV v3.1**: Apply at [PhysioNet](https://physionet.org/content/mimiciv/3.1/) → complete the required training (CITI "Data or Specimens Only Research") and sign the PhysioNet DUA. Access is granted individually; no shared credentials are permitted.
- **eICU-CRD v2.0**: Apply at [PhysioNet](https://physionet.org/content/eicu-crd/2.0/) → same CITI training + DUA process.

After approval, download the data to a local directory and pass the path via --mimic-db and --eicu-dir arguments (or set MIMIC_DUCKDB and EICU_DIR environment variables).

## Script → Table/Figure/Number Mapping

| Script | Outputs | Manuscript Location |
|--------|---------|-------------------|
| `01_extract_mimic.sql` | MIMIC CRT records + adult stays | §2.1, §2.3.1, Figure 1 |
| `02_extract_eicu.sql` | eICU CRT records + adult stays + APACHE | §2.2, §2.3.2, Figure 1 |
| `03_availability.py` | MIMIC-IV 75,421 / 94,458 = 79.85% (first 24 h) and 81,336 / 94,458 = 86.1% (all course); eICU-CRD 12,707 / 200,234 = 6.35%; gap 73.5 pp | Table 1, Figure 2A, Figure 1, §3.2 |
| `04_recording_patterns.py` | 7.0 (median 6, IQR 4–10) / 27.5 (median 13, IQR 6–29) records per stay; time windows; eICU value distribution 349,725 records (180,145 normal / 46,975 < 2 s / 44,537 > 2 s / 37,009 hands / 41,059 feet) = 271,657 state + 78,068 site | Table 1, Table 3, Figure 2B, §3.3 |
| `05_bilateral_mimic.py` | 99.6% bilateral; 99.4% agreement; κ=0.914; mean |Δt| between sides 0.12 h | Table 2, §3.7 |
| `06_site_concordance_eicu.py` | 2,155→2,147→1,906/241 (88.8%/11.2%) | §3.8, Table 1 |
| `07_selection_bias_eicu.py` | APACHE IV 60.0 vs 54.4; acute physiology score 48.5 vs 42.7; predicted ICU mortality 5.95 vs 4.90 (APACHE-based n = 11,032 / 137,127); SMD 0.21 / 0.24 / 0.04 | Table 4, §3.6 |
| `08_selection_bias_mimic.py` | Age 64.6 vs 65.5 y; male 56.4% vs 54.2%; death 11.3% vs 14.7%; LOS 3.8 vs 3.0 d | Table 4, §3.6 |
| `09_time_window_tails.py` | Pre-admission 0.7% / 1.1%; medians 0.6 / 0.3 h; >48 h = 735 / 91 (358 >7 d, 46 >30 d) | §2.5, §4.2, §4.5 |

## Environment

- **DuckDB**: Package outputs were produced and verified on DuckDB v1.5.5 (requirements-pinned).
- **Python**: 3.10+ (tested with 3.14)
- **OS**: Windows (paths use `/` for DuckDB compatibility)
- **MIMIC-IV**: v3.1 (2008–2022, 94,458 adult ICU stays)
- **eICU-CRD**: v2.0 (2014–2015, 200,234 adult ICU stays; 625 stays excluded — 530 with age < 18 and 95 with age unrecorded)

## Key Technical Notes

1. **eICU age is VARCHAR**: ages above 89 are top-coded as the literal string `> 89` and mapped to 90 for filtering (section 2.2); ages that are unrecorded do not satisfy the adult predicate and are excluded
2. **eICU CRT source**: `nurseAssessment` table (NOT `nurseCharting`)
3. **eICU state + site**: Stored as separate rows in `cellattributevalue`; state values (`normal`/`< 2 seconds`/`> 2 seconds`) and site values (`hands`/`feet`) are not mutually exclusive
4. **MIMIC bilateral**: Left (224308) and right (223951) hand itemids
5. **Time windows**: MIMIC uses first-24-hour window; eICU uses all-course (different `intime` precision)
6. **eICU record-level statistics are restricted to the adult cohort** (200,234 stays), so record-level numerators and stay-level denominators share the same population (e.g. 349,725 CRT records across 12,707 adult CRT-available stays)

## Citation

If you use this code, please cite the paper and this OSF repository.
