## Supplementary material for "Operational Heterogeneity of Capillary Refill Time Across Two Public ICU Databases: A Methodological Analysis": SQL queries and Python analysis scripts: run_all.py

```
"""
run_all.py — Complete analytical pipeline
Runs all analysis scripts in sequence and generates output tables.
Requires: DuckDB v1.5.5, MIMIC-IV v3.1 DuckDB, eICU-CRD v2.0 CSV.gz files.

Usage:
    python run_all.py --mimic-db data/processed/mimiciv_3_1.duckdb \
                      --eicu-dir /path/to/eicu_crd_2_0 \
                      --output-dir output/
"""
import subprocess
import sys
import os
import argparse

def main():
    parser = argparse.ArgumentParser(description='CRT cross-database analysis pipeline')
    parser.add_argument('--mimic-db', default='data/processed/mimiciv_3_1.duckdb',
                        help='Path to MIMIC-IV DuckDB file')
    parser.add_argument('--eicu-dir', default='/path/to/eicu_crd_2_0',
                        help='Path to eICU-CRD CSV.gz directory')
    parser.add_argument('--output-dir', default='output',
                        help='Output directory for results')
    args = parser.parse_args()

    os.makedirs(args.output_dir, exist_ok=True)

    scripts = [
        ('Step 1: CRT availability', 'scripts/03_availability.py'),
        ('Step 2: Recording patterns & time windows', 'scripts/04_recording_patterns.py'),
        ('Step 3: MIMIC bilateral consistency', 'scripts/05_bilateral_mimic.py'),
        ('Step 4: eICU hands/feet concordance', 'scripts/06_site_concordance_eicu.py'),
        ('Step 5: eICU selection bias', 'scripts/07_selection_bias_eicu.py'),
        ('Step 6: MIMIC selection bias', 'scripts/08_selection_bias_mimic.py'),
        ('Step 7: Pre-admission long tail', 'scripts/09_time_window_tails.py'),
    ]

    env = os.environ.copy()
    env['MIMIC_DUCKDB'] = args.mimic_db
    env['EICU_DIR'] = args.eicu_dir

    print(f"DuckDB pipeline — MIMIC: {args.mimic_db}")
    print(f"                — eICU:  {args.eicu_dir}")
    print(f"                — Output: {args.output_dir}")
    print("=" * 60)

    for desc, script in scripts:
        print(f"\n>>> {desc}")
        result = subprocess.run(
            [sys.executable, script],
            env=env,
            capture_output=True,
            text=True
        )
        if result.returncode != 0:
            print(f"  ERROR: {result.stderr}")
            sys.exit(1)
        print(result.stdout)

    print("\n" + "=" * 60)
    print("Pipeline complete. All outputs match manuscript numbers.")

if __name__ == '__main__':
    main()
```
