## Supplementary material for "Operational Heterogeneity of Capillary Refill Time Across Two Public ICU Databases: A Methodological Analysis": SQL queries and Python analysis scripts: 03_availability.py

```
"""
03_availability.py — Dimension 1: CRT documentation availability
Outputs: Table 1 (availability row), Figure 1 (flow diagram numbers), Figure 2A,
         and the MIMIC all-course availability reported in Section 3.2.
"""
import duckdb
import os

DB_MIMIC = os.environ.get('MIMIC_DUCKDB', 'data/processed/mimiciv_3_1.duckdb')
EICU_DIR = os.environ.get('EICU_DIR', 'E:/MIMIC/eicu_crd_2_0')

con = duckdb.connect()

# Load MIMIC (note: tables live in schema `mimic` inside the DB => mimic.mimic.<table>)
con.execute(f"ATTACH '{DB_MIMIC}' AS mimic (READ_ONLY)")
con.execute("""
CREATE TEMP VIEW adult_stays AS
SELECT ie.stay_id, ie.hadm_id, ie.subject_id, ie.intime, ie.outtime, ie.los,
       p.anchor_age, p.gender, a.deathtime
FROM mimic.mimic.icustays ie
JOIN mimic.mimic.patients p ON p.subject_id = ie.subject_id
JOIN mimic.mimic.admissions a ON a.hadm_id = ie.hadm_id
WHERE p.anchor_age >= 18;
""")

# Load eICU (self-contained)
con.execute(f"CREATE VIEW eicu_patient AS SELECT * FROM read_csv_auto('{EICU_DIR}/patient.csv.gz')")
con.execute(f"CREATE VIEW eicu_nurseAssessment AS SELECT * FROM read_csv_auto('{EICU_DIR}/nurseAssessment.csv.gz')")
con.execute("""
CREATE VIEW eicu_adult_stays AS
SELECT patientunitstayid, uniquepid,
       CASE WHEN age = '> 89' THEN 90 ELSE TRY_CAST(age AS INTEGER) END AS age_mapped, gender,
       unitdischargestatus,
       hospitaldischargestatus,
       CASE WHEN hospitaldischargestatus = 'Expired' THEN 1 ELSE 0 END AS hospital_death
FROM eicu_patient
WHERE age = '> 89' OR TRY_CAST(age AS INTEGER) >= 18;
""")
con.execute("""
CREATE VIEW eicu_crt_records AS
SELECT na.patientunitstayid, na.nurseassessoffset, na.cellattribute, na.cellattributevalue,
       CASE WHEN na.cellattributevalue IN ('normal', '< 2 seconds') THEN 'normal'
            WHEN na.cellattributevalue = '> 2 seconds' THEN 'abnormal' ELSE NULL END AS binary_state,
       CASE WHEN na.cellattributevalue = 'hands' THEN 'hands'
            WHEN na.cellattributevalue = 'feet' THEN 'feet' ELSE NULL END AS site
FROM eicu_nurseAssessment na
WHERE na.celllabel LIKE '%apillary%';
""")

# --- MIMIC: first-24h availability ---
mimic_total = con.execute("SELECT COUNT(*) FROM adult_stays").fetchone()[0]
mimic_crt_stays = con.execute("""
    SELECT COUNT(DISTINCT ce.stay_id)
    FROM mimic.mimic.chartevents ce
    JOIN adult_stays s ON ce.stay_id = s.stay_id
    WHERE ce.itemid IN (223951, 224308)
      AND ce.charttime BETWEEN s.intime AND s.intime + INTERVAL '24 hours'
""").fetchone()[0]

# --- MIMIC: all-course availability (no time restriction; reported in Section 3.2) ---
# The adult filter is vacuous in MIMIC-IV v3.1 (0 patients under 18), but is stated
# explicitly so that numerator and denominator share the same population.
mimic_crt_stays_all = con.execute("""
    SELECT COUNT(DISTINCT ce.stay_id)
    FROM mimic.mimic.chartevents ce
    JOIN adult_stays s ON ce.stay_id = s.stay_id
    WHERE ce.itemid IN (223951, 224308)
""").fetchone()[0]

# --- eICU: all-course availability (restricted to adult stays, so that the
# numerator and the 200,234-stay denominator share the same population) ---
eicu_total = con.execute("SELECT COUNT(*) FROM eicu_adult_stays").fetchone()[0]
eicu_crt_stays = con.execute("""
    SELECT COUNT(DISTINCT c.patientunitstayid)
    FROM eicu_crt_records c
    JOIN eicu_adult_stays s ON c.patientunitstayid = s.patientunitstayid
    WHERE c.binary_state IS NOT NULL OR c.site IS NOT NULL
""").fetchone()[0]

# --- Output ---
print("=== CRT Documentation Availability ===")
print(f"MIMIC-IV:  {mimic_crt_stays:,} / {mimic_total:,} = {mimic_crt_stays/mimic_total*100:.2f}% (first 24h)")
print(f"MIMIC-IV:  {mimic_crt_stays_all:,} / {mimic_total:,} = {mimic_crt_stays_all/mimic_total*100:.2f}% (all course)")
print(f"eICU-CRD:  {eicu_crt_stays:,} / {eicu_total:,} = {eicu_crt_stays/eicu_total*100:.2f}% (all course)")
print(f"Gap:       {(mimic_crt_stays/mimic_total - eicu_crt_stays/eicu_total)*100:.1f} pp")

con.close()
```
