## Supplementary material for "Operational Heterogeneity of Capillary Refill Time Across Two Public ICU Databases: A Methodological Analysis": SQL queries and Python analysis scripts: 04_recording_patterns.py

```
"""
04_recording_patterns.py — Dimension 2 & 3: Recording patterns + time windows
Outputs: Table 1 (records/stay), Table 2 (recording patterns), Table 3 (time windows), Figure 2B
"""
import duckdb
import os

DB_MIMIC = os.environ.get('MIMIC_DUCKDB', 'data/processed/mimiciv_3_1.duckdb')
EICU_DIR = os.environ.get('EICU_DIR', 'E:/MIMIC/eicu_crd_2_0')

con = duckdb.connect()
con.execute(f"ATTACH '{DB_MIMIC}' AS mimic (READ_ONLY)")

# Self-contained MIMIC view
con.execute("""
CREATE TEMP VIEW adult_stays AS
SELECT ie.stay_id, ie.hadm_id, ie.subject_id, ie.intime, ie.outtime, ie.los,
       p.anchor_age, p.gender, a.deathtime
FROM mimic.mimic.icustays ie
JOIN mimic.mimic.patients p ON p.subject_id = ie.subject_id
JOIN mimic.mimic.admissions a ON a.hadm_id = ie.hadm_id
WHERE p.anchor_age >= 18;
""")

# Self-contained eICU views
con.execute(f"CREATE VIEW eicu_patient AS SELECT * FROM read_csv_auto('{EICU_DIR}/patient.csv.gz')")
con.execute(f"CREATE VIEW eicu_nurseAssessment AS SELECT * FROM read_csv_auto('{EICU_DIR}/nurseAssessment.csv.gz')")
# eICU patient.age is VARCHAR in v2.0: ages above 89 are top-coded as the literal
# string '> 89'; unrecorded ages do not satisfy the adult predicate and are excluded.
con.execute("""
CREATE VIEW eicu_adult_stays AS
SELECT patientunitstayid, uniquepid,
       CASE WHEN age = '> 89' THEN 90 ELSE TRY_CAST(age AS INTEGER) END AS age_mapped, gender,
       unitdischargestatus,
       hospitaldischargestatus,
       CASE WHEN hospitaldischargestatus = 'Expired' THEN 1 ELSE 0 END AS hospital_death
FROM eicu_patient
WHERE age = '> 89' OR TRY_CAST(age AS INTEGER) >= 18;
""")
con.execute("""
CREATE VIEW eicu_crt_records AS
SELECT na.patientunitstayid, na.nurseassessoffset, na.cellattribute, na.cellattributevalue,
       CASE WHEN na.cellattributevalue IN ('normal', '< 2 seconds') THEN 'normal'
            WHEN na.cellattributevalue = '> 2 seconds' THEN 'abnormal' ELSE NULL END AS binary_state,
       CASE WHEN na.cellattributevalue = 'hands' THEN 'hands'
            WHEN na.cellattributevalue = 'feet' THEN 'feet' ELSE NULL END AS site
FROM eicu_nurseAssessment na
WHERE na.celllabel LIKE '%apillary%';
""")

# --- MIMIC: records per stay (first 24h) ---
mimic_rps = con.execute("""
    WITH crt AS (
        SELECT ce.stay_id, COUNT(*) AS n_records
        FROM mimic.mimic.chartevents ce
        JOIN adult_stays s ON ce.stay_id = s.stay_id
        WHERE ce.itemid IN (223951, 224308)
          AND ce.charttime BETWEEN s.intime AND s.intime + INTERVAL '24 hours'
        GROUP BY ce.stay_id
    )
    SELECT
        ROUND(AVG(n_records), 2) AS mean_records,
        ROUND(median(n_records), 1) AS median_records,
        ROUND(quantile_cont(n_records, 0.25), 1) AS q25,
        ROUND(quantile_cont(n_records, 0.75), 1) AS q75
    FROM crt
""").fetchone()

# --- eICU: records per stay (all-course, adult CRT-available stays,
# state + site rows, matching the 27.5 reported in Table 1) ---
eicu_rps = con.execute("""
    WITH crt_stays AS (
        SELECT DISTINCT patientunitstayid
        FROM eicu_crt_records
        WHERE binary_state IS NOT NULL
    ),
    adult_crt_stays AS (
        SELECT c.patientunitstayid
        FROM crt_stays c
        JOIN eicu_adult_stays s ON c.patientunitstayid = s.patientunitstayid
    ),
    crt AS (
        SELECT r.patientunitstayid, COUNT(*) AS n_records
        FROM eicu_crt_records r
        JOIN adult_crt_stays a ON r.patientunitstayid = a.patientunitstayid
        GROUP BY r.patientunitstayid
    )
    SELECT
        ROUND(AVG(n_records), 2) AS mean_records,
        ROUND(median(n_records), 1) AS median_records,
        ROUND(quantile_cont(n_records, 0.25), 1) AS q25,
        ROUND(quantile_cont(n_records, 0.75), 1) AS q75
    FROM crt
""").fetchone()

# --- MIMIC: time window distribution ---
mimic_windows = con.execute("""
    WITH crt AS (
        SELECT ce.stay_id,
               (EPOCH(ce.charttime) - EPOCH(s.intime)) / 3600.0 AS hours_from_adm
        FROM mimic.mimic.chartevents ce
        JOIN adult_stays s ON ce.stay_id = s.stay_id
        WHERE ce.itemid IN (223951, 224308)
    )
    SELECT
        CASE
            WHEN hours_from_adm < 0 THEN 'Pre-admission'
            WHEN hours_from_adm < 6 THEN '0-6h'
            WHEN hours_from_adm < 12 THEN '6-12h'
            WHEN hours_from_adm < 24 THEN '12-24h'
            ELSE '>24h'
        END AS time_window,
        COUNT(*) AS n_records,
        ROUND(COUNT(*) * 100.0 / (SELECT COUNT(*) FROM crt), 1) AS pct
    FROM crt
    GROUP BY 1
    ORDER BY MIN(hours_from_adm)
""").fetchall()

# --- eICU: time window distribution (adult cohort; same population as the 12,707
# CRT-available stays, so that record-level and stay-level denominators agree) ---
eicu_windows = con.execute("""
    WITH crt AS (
        SELECT r.patientunitstayid, r.nurseassessoffset
        FROM eicu_crt_records r
        JOIN eicu_adult_stays s ON r.patientunitstayid = s.patientunitstayid
        WHERE r.binary_state IS NOT NULL
    )
    SELECT
        CASE
            WHEN nurseassessoffset < 0 THEN 'Pre-admission'
            WHEN nurseassessoffset < 360 THEN '0-6h'
            WHEN nurseassessoffset < 720 THEN '6-12h'
            WHEN nurseassessoffset < 1440 THEN '12-24h'
            ELSE '>24h'
        END AS time_window,
        COUNT(*) AS n_records,
        ROUND(COUNT(*) * 100.0 / (SELECT COUNT(*) FROM crt), 1) AS pct
    FROM crt
    GROUP BY 1
    ORDER BY MIN(nurseassessoffset)
""").fetchall()

# --- eICU: CRT value distribution (adult cohort; Section 3.3 five categories) ---
# State values (normal / < 2 seconds / > 2 seconds) and site values (hands / feet) are
# stored as separate rows and are not mutually exclusive; percentages are of all five
# categories combined.
eicu_values = con.execute("""
    WITH crt AS (
        SELECT r.patientunitstayid, r.cellattributevalue
        FROM eicu_crt_records r
        JOIN eicu_adult_stays s ON r.patientunitstayid = s.patientunitstayid
        WHERE r.cellattributevalue IN ('normal', '< 2 seconds', '> 2 seconds', 'hands', 'feet')
    )
    SELECT cellattributevalue AS value,
           COUNT(*) AS n_records,
           COUNT(DISTINCT patientunitstayid) AS n_stays,
           ROUND(COUNT(*) * 100.0 / (SELECT COUNT(*) FROM crt), 1) AS pct
    FROM crt
    GROUP BY 1
    ORDER BY n_records DESC
""").fetchall()

print("=== Records per Stay ===")
print(f"MIMIC-IV:  mean={mimic_rps[0]}, median={mimic_rps[1]} (IQR {mimic_rps[2]}-{mimic_rps[3]})")
print(f"eICU-CRD:  mean={eicu_rps[0]}, median={eicu_rps[1]} (IQR {eicu_rps[2]}-{eicu_rps[3]})")

print("\n=== Time Window Distribution (MIMIC-IV) ===")
for row in mimic_windows:
    print(f"  {row[0]:15s}: {row[1]:>8,} records ({row[2]:5.1f}%)")

print("\n=== Time Window Distribution (eICU-CRD) ===")
for row in eicu_windows:
    print(f"  {row[0]:15s}: {row[1]:>8,} records ({row[2]:5.1f}%)")

print("\n=== CRT Value Distribution (eICU-CRD, adult cohort) ===")
tot_rec = sum(r[1] for r in eicu_values)
for row in eicu_values:
    print(f"  {row[0]:14s}: {row[1]:>7,} records across {row[2]:>6,} stays ({row[3]:5.1f}%)")
print(f"  {'TOTAL':14s}: {tot_rec:>7,} records")
state_rec = sum(r[1] for r in eicu_values if r[0] in ('normal', '< 2 seconds', '> 2 seconds'))
site_rec = sum(r[1] for r in eicu_values if r[0] in ('hands', 'feet'))
print(f"  state values  : {state_rec:>7,} records")
print(f"  site values   : {site_rec:>7,} records")

con.close()
```
