## Supplementary material for "Operational Heterogeneity of Capillary Refill Time Across Two Public ICU Databases: A Methodological Analysis": SQL queries and Python analysis scripts: 05_bilateral_mimic.py

```
"""
05_bilateral_mimic.py — Dimension 4a: MIMIC-IV bilateral (left/right) consistency
Outputs: Table 2 (bilateral rows), Table 1 (bilateral / percent agreement / Cohen's kappa),
         discordant breakdown, timestamp concordance.
"""
import duckdb
import os

DB_MIMIC = os.environ.get('MIMIC_DUCKDB', 'data/processed/mimiciv_3_1.duckdb')
con = duckdb.connect()
con.execute(f"ATTACH '{DB_MIMIC}' AS mimic (READ_ONLY)")

# Self-contained adult-stays view (no dependency on 01_extract_mimic.sql)
con.execute("""
CREATE TEMP VIEW adult_stays AS
SELECT ie.stay_id, ie.hadm_id, ie.subject_id, ie.intime, ie.outtime, ie.los,
       p.anchor_age, p.gender, a.deathtime
FROM mimic.mimic.icustays ie
JOIN mimic.mimic.patients p ON p.subject_id = ie.subject_id
JOIN mimic.mimic.admissions a ON a.hadm_id = ie.hadm_id
WHERE p.anchor_age >= 18;
""")

# --- Bilateral availability ---
bilateral_stats = con.execute("""
    WITH crt AS (
        SELECT DISTINCT ce.stay_id, ce.itemid
        FROM mimic.mimic.chartevents ce
        JOIN adult_stays s ON ce.stay_id = s.stay_id
        WHERE ce.itemid IN (223951, 224308)
          AND ce.charttime BETWEEN s.intime AND s.intime + INTERVAL '24 hours'
    ),
    per_stay AS (
        SELECT stay_id, COUNT(DISTINCT itemid) AS n_sides
        FROM crt
        GROUP BY stay_id
    )
    SELECT
        COUNT(*) AS total_crt_stays,
        SUM(CASE WHEN n_sides = 2 THEN 1 ELSE 0 END) AS bilateral_stays,
        ROUND(SUM(CASE WHEN n_sides = 2 THEN 100.0 ELSE 0 END) / COUNT(*), 1) AS bilateral_pct
    FROM per_stay
""").fetchone()

# --- Agreement analysis (most recent record per side per stay) ---
agreement = con.execute("""
    WITH latest_per_side AS (
        SELECT
            ce.stay_id,
            ce.itemid,
            CASE WHEN ce.itemid = 223951 THEN 'right' ELSE 'left' END AS side,
            CASE WHEN lower(ce.value) LIKE '%abnormal%' THEN 'abnormal' WHEN lower(ce.value) LIKE '%normal%' THEN 'normal' ELSE 'other' END AS binary_state,
            ROW_NUMBER() OVER (
                PARTITION BY ce.stay_id, ce.itemid
                ORDER BY ce.charttime DESC
            ) AS rn
        FROM mimic.mimic.chartevents ce
        JOIN adult_stays s ON ce.stay_id = s.stay_id
        WHERE ce.itemid IN (223951, 224308)
          AND ce.charttime BETWEEN s.intime AND s.intime + INTERVAL '24 hours'
    ),
    paired AS (
        SELECT
            r.stay_id,
            r.binary_state AS right_state,
            l.binary_state AS left_state,
            CASE WHEN r.binary_state = l.binary_state THEN 'concordant' ELSE 'discordant' END AS agreement
        FROM latest_per_side r
        JOIN latest_per_side l ON r.stay_id = l.stay_id
        WHERE r.side = 'right' AND l.side = 'left' AND r.rn = 1 AND l.rn = 1
    )
    SELECT
        agreement,
        COUNT(*) AS n_stays,
        ROUND(COUNT(*) * 100.0 / (SELECT COUNT(*) FROM paired), 1) AS pct
    FROM paired
    GROUP BY agreement
""").fetchall()

# --- Discordant breakdown ---
discordant = con.execute("""
    WITH latest_per_side AS (
        SELECT
            ce.stay_id,
            CASE WHEN ce.itemid = 223951 THEN 'right' ELSE 'left' END AS side,
            CASE WHEN lower(ce.value) LIKE '%abnormal%' THEN 'abnormal' WHEN lower(ce.value) LIKE '%normal%' THEN 'normal' ELSE 'other' END AS binary_state,
            ROW_NUMBER() OVER (
                PARTITION BY ce.stay_id, ce.itemid
                ORDER BY ce.charttime DESC
            ) AS rn
        FROM mimic.mimic.chartevents ce
        JOIN adult_stays s ON ce.stay_id = s.stay_id
        WHERE ce.itemid IN (223951, 224308)
          AND ce.charttime BETWEEN s.intime AND s.intime + INTERVAL '24 hours'
    ),
    paired AS (
        SELECT
            r.stay_id,
            r.binary_state AS right_state,
            l.binary_state AS left_state
        FROM latest_per_side r
        JOIN latest_per_side l ON r.stay_id = l.stay_id
        WHERE r.side = 'right' AND l.side = 'left' AND r.rn = 1 AND l.rn = 1
          AND r.binary_state != l.binary_state
    )
    SELECT right_state, left_state, COUNT(*) AS n
    FROM paired
    GROUP BY right_state, left_state
""").fetchall()

# --- Cohen's kappa over the paired 2x2 table (right x left) ---
kappa_2x2 = con.execute("""
    WITH latest_per_side AS (
        SELECT
            ce.stay_id,
            CASE WHEN ce.itemid = 223951 THEN 'right' ELSE 'left' END AS side,
            CASE WHEN lower(ce.value) LIKE '%abnormal%' THEN 'abnormal' WHEN lower(ce.value) LIKE '%normal%' THEN 'normal' ELSE 'other' END AS binary_state,
            ROW_NUMBER() OVER (
                PARTITION BY ce.stay_id, ce.itemid
                ORDER BY ce.charttime DESC
            ) AS rn
        FROM mimic.mimic.chartevents ce
        JOIN adult_stays s ON ce.stay_id = s.stay_id
        WHERE ce.itemid IN (223951, 224308)
          AND ce.charttime BETWEEN s.intime AND s.intime + INTERVAL '24 hours'
    ),
    paired AS (
        SELECT r.binary_state AS right_state, l.binary_state AS left_state
        FROM latest_per_side r
        JOIN latest_per_side l ON r.stay_id = l.stay_id
        WHERE r.side = 'right' AND l.side = 'left' AND r.rn = 1 AND l.rn = 1
    )
    SELECT
        SUM(CASE WHEN right_state = 'normal'   AND left_state = 'normal'   THEN 1 ELSE 0 END) AS nn,
        SUM(CASE WHEN right_state = 'normal'   AND left_state = 'abnormal' THEN 1 ELSE 0 END) AS na,
        SUM(CASE WHEN right_state = 'abnormal' AND left_state = 'normal'   THEN 1 ELSE 0 END) AS an,
        SUM(CASE WHEN right_state = 'abnormal' AND left_state = 'abnormal' THEN 1 ELSE 0 END) AS aa,
        COUNT(*) AS n
    FROM paired
""").fetchone()

nn, na, an, aa, n_paired = kappa_2x2
po = (nn + aa) / n_paired
pe = ((nn + na) * (nn + an) + (an + aa) * (na + aa)) / (n_paired ** 2)
kappa = (po - pe) / (1 - pe) if pe < 1 else float('nan')

# --- Timestamp concordance ---
timestamp_conc = con.execute("""
    WITH latest_per_side AS (
        SELECT
            ce.stay_id,
            CASE WHEN ce.itemid = 223951 THEN 'right' ELSE 'left' END AS side,
            ce.charttime,
            ROW_NUMBER() OVER (
                PARTITION BY ce.stay_id, ce.itemid
                ORDER BY ce.charttime DESC
            ) AS rn
        FROM mimic.mimic.chartevents ce
        JOIN adult_stays s ON ce.stay_id = s.stay_id
        WHERE ce.itemid IN (223951, 224308)
          AND ce.charttime BETWEEN s.intime AND s.intime + INTERVAL '24 hours'
    ),
    paired AS (
        SELECT
            r.stay_id,
            CASE WHEN r.charttime = l.charttime THEN 'same' ELSE 'different' END AS ts_match
        FROM latest_per_side r
        JOIN latest_per_side l ON r.stay_id = l.stay_id
        WHERE r.side = 'right' AND l.side = 'left' AND r.rn = 1 AND l.rn = 1
    )
    SELECT ts_match, COUNT(*) AS n, ROUND(COUNT(*) * 100.0 / (SELECT COUNT(*) FROM paired), 1) AS pct
    FROM paired GROUP BY ts_match
""").fetchall()

# --- Timestamp difference statistics over the paired stays (§3.7: mean |Δt| = 0.12 h) ---
ts_stats = con.execute("""
    WITH latest_per_side AS (
        SELECT
            ce.stay_id,
            CASE WHEN ce.itemid = 223951 THEN 'right' ELSE 'left' END AS side,
            ce.charttime,
            ROW_NUMBER() OVER (
                PARTITION BY ce.stay_id, ce.itemid
                ORDER BY ce.charttime DESC
            ) AS rn
        FROM mimic.mimic.chartevents ce
        JOIN adult_stays s ON ce.stay_id = s.stay_id
        WHERE ce.itemid IN (223951, 224308)
          AND ce.charttime BETWEEN s.intime AND s.intime + INTERVAL '24 hours'
    ),
    paired AS (
        SELECT ABS(EXTRACT(EPOCH FROM r.charttime) - EXTRACT(EPOCH FROM l.charttime)) / 3600.0 AS dt_hours
        FROM latest_per_side r
        JOIN latest_per_side l ON r.stay_id = l.stay_id
        WHERE r.side = 'right' AND l.side = 'left' AND r.rn = 1 AND l.rn = 1
    )
    SELECT ROUND(AVG(dt_hours), 2) AS mean_h,
           ROUND(MEDIAN(dt_hours), 2) AS median_h,
           ROUND(QUANTILE_CONT(dt_hours, 0.75), 2) AS p75_h
    FROM paired
""").fetchone()

print("=== MIMIC-IV Bilateral Consistency ===")
print(f"Bilateral availability: {bilateral_stats[1]:,} / {bilateral_stats[0]:,} = {bilateral_stats[2]}%")
print(f"\nAgreement:")
for row in agreement:
    print(f"  {row[0]}: {row[1]:,} ({row[2]}%)")
print(f"\nDiscordant breakdown:")
for row in discordant:
    print(f"  R={row[0]}, L={row[1]}: {row[2]}")

print(f"\n2x2 (right x left), paired most-recent assessments: n = {n_paired:,}")
print(f"  R-normal/L-normal   = {nn:,}")
print(f"  R-normal/L-abnormal = {na:,}")
print(f"  R-abnormal/L-normal = {an:,}")
print(f"  R-abnormal/L-abnormal = {aa:,}")
print(f"Percent agreement (Po) = {po * 100:.1f}%")
print(f"Expected agreement (Pe) = {pe * 100:.1f}%")
print(f"Cohen's kappa = {kappa:.3f}")

print(f"\nTimestamp concordance:")
for row in timestamp_conc:
    print(f"  {row[0]}: {row[1]:,} ({row[2]}%)")
print(f"Mean |Δt| between sides (paired stays): {ts_stats[0]} h "
      f"(median {ts_stats[1]} h, p75 {ts_stats[2]} h)")

con.close()
```
