## Supplementary material for "Operational Heterogeneity of Capillary Refill Time Across Two Public ICU Databases: A Methodological Analysis": SQL queries and Python analysis scripts: 06_site_concordance_eicu.py

```
"""
06_site_concordance_eicu.py — Dimension 4b: eICU-CRD hands/feet site concordance
Outputs: Table 1 (hands/feet rows), section 3.8 numbers.
All record-level statistics are restricted to the adult cohort so that they share the
same population as the 12,707 CRT-available adult stays and the 200,234-stay denominator.
"""
import duckdb
import os

EICU_DIR = os.environ.get('EICU_DIR', 'E:/MIMIC/eicu_crd_2_0')
con = duckdb.connect()

for tbl in ['patient', 'nurseAssessment']:
    con.execute(f"CREATE OR REPLACE VIEW eicu_{tbl} AS SELECT * FROM read_csv_auto('{EICU_DIR}/{tbl}.csv.gz')")

con.execute("""
CREATE VIEW eicu_adult_stays AS
SELECT patientunitstayid
FROM eicu_patient
WHERE age = '> 89' OR TRY_CAST(age AS INTEGER) >= 18;
""")
con.execute("""
CREATE VIEW eicu_crt_adult AS
SELECT na.patientunitstayid, na.nurseassessoffset, na.cellattributevalue
FROM eicu_nurseAssessment na
JOIN eicu_adult_stays s ON na.patientunitstayid = s.patientunitstayid
WHERE na.celllabel LIKE '%apillary%';
""")

# --- Hands/feet availability ---
site_counts = con.execute("""
    WITH crt AS (
        SELECT patientunitstayid, cellattributevalue,
               CASE WHEN cellattributevalue = 'hands' THEN 'hands'
                    WHEN cellattributevalue = 'feet' THEN 'feet' END AS site
        FROM eicu_crt_adult
        WHERE cellattributevalue IN ('hands', 'feet')
    )
    SELECT site, COUNT(DISTINCT patientunitstayid) AS n_stays
    FROM crt GROUP BY site
""").fetchall()

both_sites = con.execute("""
    WITH crt AS (
        SELECT patientunitstayid,
               CASE WHEN cellattributevalue = 'hands' THEN 1 END AS is_hands,
               CASE WHEN cellattributevalue = 'feet' THEN 1 END AS is_feet
        FROM eicu_crt_adult
    )
    SELECT COUNT(*) AS n_both
    FROM (
        SELECT patientunitstayid
        FROM crt
        GROUP BY patientunitstayid
        HAVING SUM(is_hands) > 0 AND SUM(is_feet) > 0
    )
""").fetchone()[0]

# --- Concordance analysis (same method as paper) ---
concordance = con.execute("""
    WITH crt_all AS (
        SELECT patientunitstayid, nurseassessoffset, cellattributevalue,
               CASE WHEN cellattributevalue IN ('normal', '< 2 seconds') THEN 'normal'
                    WHEN cellattributevalue = '> 2 seconds' THEN 'abnormal' END AS binary_state,
               CASE WHEN cellattributevalue = 'hands' THEN 'hands'
                    WHEN cellattributevalue = 'feet' THEN 'feet' END AS site
        FROM eicu_crt_adult
    ),
    offset_states AS (
        SELECT patientunitstayid, nurseassessoffset, MAX(binary_state) AS state_value
        FROM crt_all WHERE binary_state IS NOT NULL
        GROUP BY patientunitstayid, nurseassessoffset
    ),
    offset_sites AS (
        SELECT patientunitstayid, nurseassessoffset,
               MAX(CASE WHEN site = 'hands' THEN 1 ELSE 0 END) AS has_hands,
               MAX(CASE WHEN site = 'feet' THEN 1 ELSE 0 END) AS has_feet
        FROM crt_all WHERE site IS NOT NULL
        GROUP BY patientunitstayid, nurseassessoffset
    ),
    site_states AS (
        SELECT os.patientunitstayid, os.nurseassessoffset, os.state_value,
               oss.has_hands, oss.has_feet
        FROM offset_states os
        JOIN offset_sites oss ON os.patientunitstayid = oss.patientunitstayid
            AND os.nurseassessoffset = oss.nurseassessoffset
    ),
    stays_both AS (
        SELECT patientunitstayid FROM crt_all WHERE site IS NOT NULL
        GROUP BY patientunitstayid HAVING COUNT(DISTINCT site) = 2
    ),
    hands_latest AS (
        SELECT patientunitstayid, state_value,
               ROW_NUMBER() OVER (PARTITION BY patientunitstayid ORDER BY nurseassessoffset DESC) AS rn
        FROM site_states WHERE has_hands = 1 AND state_value IS NOT NULL
    ),
    feet_latest AS (
        SELECT patientunitstayid, state_value,
               ROW_NUMBER() OVER (PARTITION BY patientunitstayid ORDER BY nurseassessoffset DESC) AS rn
        FROM site_states WHERE has_feet = 1 AND state_value IS NOT NULL
    )
    SELECT h.state_value AS hands_state, f.state_value AS feet_state,
           CASE WHEN h.state_value = f.state_value THEN 'concordant' ELSE 'discordant' END AS agreement,
           COUNT(*) AS n_stays
    FROM stays_both sb
    JOIN hands_latest h ON sb.patientunitstayid = h.patientunitstayid AND h.rn = 1
    JOIN feet_latest f ON sb.patientunitstayid = f.patientunitstayid AND f.rn = 1
    GROUP BY h.state_value, f.state_value
""").fetchall()

total_valid = sum(r[3] for r in concordance)

print("=== eICU-CRD Hands/Feet Site Concordance (adult cohort) ===")
site_map = dict(site_counts)
hands_stays = site_map.get('hands', 0)
feet_stays = site_map.get('feet', 0)
print(f"Hands sites: {hands_stays:,} stays")
print(f"Feet sites:  {feet_stays:,} stays")
print(f"Both sites:  {both_sites:,} stays")
print(f"Hands only:  {hands_stays - both_sites:,} stays")
print(f"Feet only:   {feet_stays - both_sites:,} stays")
print(f"\nValid pairs (with state at both sites): {total_valid:,}")
for row in concordance:
    pct = row[3] / total_valid * 100 if total_valid > 0 else 0
    print(f"  hands={row[0]}, feet={row[1]}: {row[3]:,} ({pct:.1f}%) [{row[2]}]")

concordant = sum(r[3] for r in concordance if r[2] == 'concordant')
discordant = sum(r[3] for r in concordance if r[2] == 'discordant')
print(f"\nConcordant: {concordant:,} ({concordant/total_valid*100:.1f}%)")
print(f"Discordant: {discordant:,} ({discordant/total_valid*100:.1f}%)")

# Directional breakdown of the discordant pairs (paper §3.8)
for hs, fs in (('normal', 'abnormal'), ('abnormal', 'normal')):
    n = sum(r[3] for r in concordance if r[0] == hs and r[1] == fs)
    print(f"  hands={hs}, feet={fs}: {n:,} ({n/total_valid*100:.1f}%)")

con.close()
```
