## Supplementary material for "Operational Heterogeneity of Capillary Refill Time Across Two Public ICU Databases: A Methodological Analysis": SQL queries and Python analysis scripts: 07_selection_bias_eicu.py

```
"""
07_selection_bias_eicu.py — Dimension 5a: eICU-CRD selection bias (APACHE IV-based)
Outputs: Table 4 (eICU columns), SMD calculation for APACHE IV score, acute physiology score,
         and predicted ICU mortality, plus the denominators of the three APACHE-based rows.
"""
import duckdb
import os
import math

EICU_DIR = os.environ.get('EICU_DIR', 'E:/MIMIC/eicu_crd_2_0')
con = duckdb.connect()

# Self-contained eICU views
con.execute(f"CREATE VIEW eicu_patient AS SELECT * FROM read_csv_auto('{EICU_DIR}/patient.csv.gz')")
con.execute(f"CREATE VIEW eicu_nurseAssessment AS SELECT * FROM read_csv_auto('{EICU_DIR}/nurseAssessment.csv.gz')")
con.execute(f"CREATE VIEW eicu_apachePatientResult AS SELECT * FROM read_csv_auto('{EICU_DIR}/apachePatientResult.csv.gz')")
con.execute("""
CREATE VIEW eicu_adult_stays AS
SELECT patientunitstayid, uniquepid,
       CASE WHEN age = '> 89' THEN 90 ELSE TRY_CAST(age AS INTEGER) END AS age_mapped, gender,
       unitdischargestatus,
       hospitaldischargestatus,
       CASE WHEN hospitaldischargestatus = 'Expired' THEN 1 ELSE 0 END AS hospital_death
FROM eicu_patient
WHERE age = '> 89' OR TRY_CAST(age AS INTEGER) >= 18;
""")
con.execute("""
CREATE VIEW eicu_crt_records AS
SELECT na.patientunitstayid, na.nurseassessoffset, na.cellattribute, na.cellattributevalue,
       CASE WHEN na.cellattributevalue IN ('normal', '< 2 seconds') THEN 'normal'
            WHEN na.cellattributevalue = '> 2 seconds' THEN 'abnormal' ELSE NULL END AS binary_state,
       CASE WHEN na.cellattributevalue = 'hands' THEN 'hands'
            WHEN na.cellattributevalue = 'feet' THEN 'feet' ELSE NULL END AS site
FROM eicu_nurseAssessment na
WHERE na.celllabel LIKE '%apillary%';
""")

# --- CRT-available vs CRT-unavailable comparison ---
# NOTE: apacheversion is fixed to 'IV'. The table also contains an 'IVa' variant
# (148,532 rows each); pooling both duplicates rows and distorts every denominator.
result = con.execute("""
    WITH crt_available AS (
        SELECT DISTINCT patientunitstayid
        FROM eicu_crt_records
        WHERE binary_state IS NOT NULL OR site IS NOT NULL
    ),
    apache AS (
        SELECT patientunitstayid,
               apachescore AS apache_iv_score,
               acutephysiologyscore AS aps,
               predictedicumortality AS pred_icu_mort
        FROM eicu_apachePatientResult
        WHERE apacheversion = 'IV'
    ),
    combined AS (
        SELECT
            s.patientunitstayid,
            CASE WHEN ca.patientunitstayid IS NOT NULL THEN 'CRT-available' ELSE 'CRT-unavailable' END AS group_name,
            s.age_mapped AS age,
            s.gender,
            s.hospital_death
        FROM eicu_adult_stays s
        LEFT JOIN crt_available ca ON s.patientunitstayid = ca.patientunitstayid
    )
    SELECT
        c.group_name,
        COUNT(*) AS n,
        COUNT(a.apache_iv_score) AS n_apache,
        ROUND(AVG(c.age), 1) AS mean_age,
        ROUND(SUM(CASE WHEN c.gender = 'Male' THEN 100.0 ELSE 0 END) / COUNT(*), 1) AS male_pct,
        ROUND(AVG(a.apache_iv_score), 1) AS mean_apache,
        ROUND(STDDEV(a.apache_iv_score), 1) AS sd_apache,
        AVG(a.apache_iv_score) AS apache_mean_raw,
        STDDEV(a.apache_iv_score) AS apache_sd_raw,
        ROUND(AVG(a.aps), 1) AS mean_aps,
        ROUND(STDDEV(a.aps), 1) AS sd_aps,
        AVG(a.aps) AS aps_mean_raw,
        STDDEV(a.aps) AS aps_sd_raw,
        ROUND(AVG(a.pred_icu_mort) * 100, 2) AS mean_pred_pct,
        ROUND(STDDEV(a.pred_icu_mort) * 100, 2) AS sd_pred_pct,
        AVG(a.pred_icu_mort) AS pred_mean_raw,
        STDDEV(a.pred_icu_mort) AS pred_sd_raw,
        ROUND(SUM(CASE WHEN c.hospital_death = 1 THEN 100.0 ELSE 0 END) / COUNT(*), 1) AS death_pct
    FROM combined c
    LEFT JOIN apache a ON c.patientunitstayid = a.patientunitstayid
    GROUP BY c.group_name
    ORDER BY c.group_name
""").fetchall()

# Standardized mean differences. Uses the unrounded group means/SDs and the APACHE-subset
# denominators: computing the SMD from the 1-decimal display values inflates it
# (5.6/26.03 = 0.22) relative to the correct 5.5538/26.0781 = 0.21 reported in the manuscript.
def smd(idx_mean, idx_sd):
    if len(result) != 2:
        return None
    n1, m1, sd1 = result[0][2], result[0][idx_mean], result[0][idx_sd]
    n2, m2, sd2 = result[1][2], result[1][idx_mean], result[1][idx_sd]
    if None in (m1, sd1, m2, sd2):
        return None
    pooled_sd = math.sqrt(((n1 - 1) * sd1**2 + (n2 - 1) * sd2**2) / (n1 + n2 - 2))
    return abs(m1 - m2) / pooled_sd if pooled_sd > 0 else 0

# (label, raw mean idx, raw sd idx, display mean idx, display sd idx, display decimals)
MEASURES = [
    ("APACHE IV score", 7, 8, 5, 6, 1),
    ("Acute physiology score", 11, 12, 9, 10, 1),
    ("Predicted ICU mortality", 15, 16, 13, 14, 2),
]

print("=== eICU-CRD Selection Bias (APACHE version 'IV') ===")
for row in result:
    print(f"\n{row[0]}: n = {row[1]:,} (APACHE-based rows: n = {row[2]:,})")
    print(f"   age {row[3]} y | male {row[4]}% | hospital mortality {row[17]}%")
    for label, _, _, mi, si, dp in MEASURES:
        print(f"   {label:24s} {row[mi]:.{dp}f} (SD {row[si]:.{dp}f})")

print("\n--- Denominators of the three APACHE-based rows ---")
for row in result:
    print(f"   {row[0]}: n = {row[2]:,} of {row[1]:,} stays ({(row[2]/row[1]*100):.1f}%)")

print("\n--- SMD (computed from unrounded means/SDs) ---")
for label, mi, si, _, _, _ in MEASURES:
    s = smd(mi, si)
    print(f"   SMD ({(label):24s}) = {s:.2f}" if s is not None else f"   SMD ({label}): n/a")

con.close()
```
