## Supplementary material for "Operational Heterogeneity of Capillary Refill Time Across Two Public ICU Databases: A Methodological Analysis": SQL queries and Python analysis scripts: 08_selection_bias_mimic.py

```
"""
08_selection_bias_mimic.py — Dimension 5b: MIMIC-IV selection bias (proxy measures)
Outputs: Table 4 (MIMIC columns), section 3.6 numbers
Note: APACHE IV / SOFA not systematically available in chartevents;
      hospital mortality and ICU LOS used as proxy severity measures.
"""
import duckdb
import os

DB_MIMIC = os.environ.get('MIMIC_DUCKDB', 'data/processed/mimiciv_3_1.duckdb')
con = duckdb.connect()
con.execute(f"ATTACH '{DB_MIMIC}' AS mimic (READ_ONLY)")

result = con.execute("""
    WITH crt_available AS (
        SELECT DISTINCT ce.stay_id
        FROM mimic.mimic.chartevents ce
        JOIN mimic.mimic.icustays s ON ce.stay_id = s.stay_id
        WHERE ce.itemid IN (223951, 224308)
          AND ce.charttime BETWEEN s.intime AND s.intime + INTERVAL '24 hours'
    )
    SELECT
        CASE WHEN ca.stay_id IS NOT NULL THEN 'CRT-available' ELSE 'CRT-unavailable' END AS group_name,
        COUNT(*) AS n,
        ROUND(AVG(p.anchor_age + (EXTRACT(YEAR FROM ic.intime) - p.anchor_year)), 1) AS mean_age,
        ROUND(SUM(CASE WHEN p.gender = 'M' THEN 100.0 ELSE 0 END) / COUNT(*), 1) AS male_pct,
        ROUND(SUM(CASE WHEN a.deathtime IS NOT NULL THEN 100.0 ELSE 0 END) / COUNT(*), 1) AS death_pct,
        ROUND(AVG(ic.los), 1) AS mean_los
    FROM mimic.mimic.icustays ic
    JOIN mimic.mimic.patients p ON ic.subject_id = p.subject_id
    JOIN mimic.mimic.admissions a ON ic.hadm_id = a.hadm_id
    LEFT JOIN crt_available ca ON ic.stay_id = ca.stay_id
    GROUP BY (ca.stay_id IS NOT NULL)
    ORDER BY (ca.stay_id IS NOT NULL)
""").fetchall()

print("=== MIMIC-IV Selection Bias (proxy measures, first 24 h) ===")
print(f"{'Group':<20s} {'n':>8s} {'Age':>6s} {'Male%':>6s} {'Death%':>7s} {'LOS':>5s}")
print("-" * 60)
for row in result:
    print(f"{row[0]:<20s} {row[1]:>8,} {row[2]:>6} {row[3]:>5}% {row[4]:>6}% {row[5]:>5}d")

# Map by group name (result order is not guaranteed)
stats = {row[0]: row for row in result}
avail = stats['CRT-available']
unavail = stats['CRT-unavailable']
print(f"\nDirection: CRT-available has {'lower' if avail[4] < unavail[4] else 'higher'} mortality "
      f"({avail[4]}% vs {unavail[4]}%) and {'longer' if avail[5] > unavail[5] else 'shorter'} LOS "
      f"({avail[5]} vs {unavail[5]} days)")
print("Interpretation: CRT documentation reflects routine practice in longer-staying patients")

con.close()
```
