## Supplementary material for "Operational Heterogeneity of Capillary Refill Time Across Two Public ICU Databases: A Methodological Analysis": SQL queries and Python analysis scripts: 09_time_window_tails.py

```
"""
09_time_window_tails.py — Dimension 3b: pre-admission CRT records and their long tail
Outputs: the 2.5 / 4.2 numbers (pre-admission shares, medians, >48 h / >7 d / >30 d counts).
"""
import duckdb
import os

DB_MIMIC = os.environ.get('MIMIC_DUCKDB', 'data/processed/mimiciv_3_1.duckdb')
EICU_DIR = os.environ.get('EICU_DIR', 'E:/MIMIC/eicu_crd_2_0')

con = duckdb.connect()
con.execute(f"ATTACH '{DB_MIMIC}' AS mimic (READ_ONLY)")

# MIMIC-IV: hours from icustays.intime (EPOCH() returns seconds, hence /3600)
m_total, m_pre, m_med, m_48, m_7d, m_30d, m_min = con.execute("""
    WITH r AS (
        SELECT (EPOCH(ce.charttime) - EPOCH(ic.intime))/3600.0 AS h
        FROM mimic.mimic.chartevents ce
        JOIN mimic.mimic.icustays ic ON ce.stay_id = ic.stay_id
        JOIN mimic.mimic.patients p ON ic.subject_id = p.subject_id
        WHERE ce.itemid IN (223951, 224308) AND p.anchor_age >= 18
    )
    SELECT COUNT(*),
           COUNT(*) FILTER (WHERE h < 0),
           ROUND(-QUANTILE_CONT(h, 0.5) FILTER (WHERE h < 0), 1),
           COUNT(*) FILTER (WHERE h < -48),
           COUNT(*) FILTER (WHERE h < -168),
           COUNT(*) FILTER (WHERE h < -720),
           ROUND(-MIN(h), 1)
    FROM r
""").fetchone()

# eICU-CRD: native nurseassessoffset (minutes from unit admission) — no intime join exists here
e_total, e_pre, e_med, e_48, e_min = con.execute(f"""
    WITH r AS (
        SELECT nurseassessoffset/60.0 AS h
        FROM read_csv_auto('{EICU_DIR}/nurseAssessment.csv.gz')
        WHERE celllabel LIKE '%apillary%'
          AND cellattributevalue IN ('normal', '< 2 seconds', '> 2 seconds')
    )
    SELECT COUNT(*),
           COUNT(*) FILTER (WHERE h < 0),
           ROUND(-QUANTILE_CONT(h, 0.5) FILTER (WHERE h < 0), 1),
           COUNT(*) FILTER (WHERE h < -48),
           ROUND(-MIN(h), 1)
    FROM r
""").fetchone()

print("=== Pre-admission CRT records and their long tail ===")
print(f"MIMIC-IV pre-admission: {m_pre:,} / {m_total:,} ({m_pre/m_total*100:.1f}%); "
      f"median {m_med} h before ICU admission; >48 h = {m_48:,}, >7 d = {m_7d:,}, "
      f">30 d = {m_30d:,}; earliest {m_min:,.1f} h (~{m_min/24:.0f} d)")
print(f"eICU-CRD pre-admission: {e_pre:,} / {e_total:,} ({e_pre/e_total*100:.1f}%); "
      f"median {e_med} h before unit admission; >48 h = {e_48:,}; earliest {e_min:,.1f} h "
      f"(~{e_min/24:.0f} d)")
con.close()
```
