## Supplementary material for "Operational Heterogeneity of Capillary Refill Time Across Two Public ICU Databases: A Methodological Analysis": SQL queries and Python analysis scripts: 01_extract_mimic.sql

```
-- =============================================================================
-- 01_extract_mimic.sql
-- MIMIC-IV v3.1 CRT extraction pipeline
-- Database: DuckDB v1.5.5, local file mimiciv_3_1.duckdb
-- Outputs: mimic_crt_records.csv, mimic_adult_stays.csv
-- RUN MODE: execute with a DIRECT connection to the DuckDB file
--   (e.g. `duckdb mimiciv_3_1.duckdb < 01_extract_mimic.sql`), NOT via ATTACH.
--   The single-prefix `mimic.<table>` references resolve to schema `mimic`
--   only under a direct connection; scripts 03-08 instead ATTACH the file
--   and therefore use the two-part `mimic.mimic.<table>` form.
-- =============================================================================

-- Step 1: Adult ICU stays (age >= 18 verified: 0 of 364,627 patients with anchor_age < 18)
CREATE OR REPLACE VIEW mimic_adult_stays AS
SELECT
    ie.stay_id,
    ie.hadm_id,
    ie.subject_id,
    ie.intime,
    ie.outtime,
    ie.los,
    p.anchor_age,
    p.gender,
    a.deathtime,
    CASE WHEN a.deathtime IS NOT NULL THEN 1 ELSE 0 END AS hospital_death
FROM mimic.icustays ie
JOIN mimic.patients p
    ON p.subject_id = ie.subject_id
JOIN mimic.admissions a
    ON a.hadm_id = ie.hadm_id
WHERE p.anchor_age >= 18;

-- Step 2: CRT records from chartevents
-- itemid 223951 = Capillary Refill Time [Right Hand]
-- itemid 224308 = Capillary Refill Time [Left Hand]
-- Values: "Normal <3 Seconds" / "Abnormal >3 Seconds"
CREATE OR REPLACE VIEW mimic_crt_records AS
SELECT
    ce.stay_id,
    ce.charttime,
    ce.itemid,
    CASE ce.itemid
        WHEN 223951 THEN 'right'
        WHEN 224308 THEN 'left'
    END AS side,
    ce.value AS raw_value,
    CASE
        WHEN lower(ce.value) LIKE '%abnormal%' THEN 'abnormal'
        WHEN lower(ce.value) LIKE '%normal%' THEN 'normal'
        ELSE ce.value
    END AS binary_state,
    -- Hours from ICU admission
    ROUND(
        (JULIANDAY(ce.charttime) - JULIANDAY(s.intime)) * 24,
        2
    ) AS hours_from_admission
FROM mimic.chartevents ce
JOIN mimic_adult_stays s
    ON ce.stay_id = s.stay_id
WHERE ce.itemid IN (223951, 224308);

-- Step 3: Export for analysis
COPY (
    SELECT * FROM mimic_adult_stays
) TO 'output/mimic_adult_stays.csv' (HEADER, DELIMITER ',');

COPY (
    SELECT * FROM mimic_crt_records
) TO 'output/mimic_crt_records.csv' (HEADER, DELIMITER ',');
```
