## Supplementary material for "Operational Heterogeneity of Capillary Refill Time Across Two Public ICU Databases: A Methodological Analysis": SQL queries and Python analysis scripts: 02_extract_eicu.sql

```
-- =============================================================================
-- 02_extract_eicu.sql
-- eICU-CRD v2.0 CRT extraction pipeline
-- Database: DuckDB v1.5.5, views over CSV.gz in eicu_crd_2_0/
-- Outputs: eicu_crt_records.csv, eicu_adult_stays.csv
-- =============================================================================

-- Create views over raw CSV.gz files
CREATE OR REPLACE VIEW eicu_patient AS
SELECT * FROM read_csv_auto('DATA_DIR/eicu_crd_2_0/patient.csv.gz');

CREATE OR REPLACE VIEW eicu_nurseAssessment AS
SELECT * FROM read_csv_auto('DATA_DIR/eicu_crd_2_0/nurseAssessment.csv.gz');

CREATE OR REPLACE VIEW eicu_apachePatientResult AS
SELECT * FROM read_csv_auto('DATA_DIR/eicu_crd_2_0/apachePatientResult.csv.gz');

-- Step 1: Adult ICU stays
-- eICU-CRD v2.0 stores patient.age as VARCHAR: ages above 89 are top-coded as the
-- literal string '> 89' (7,081 of 200,859 rows), and unrecorded ages are empty.
-- A numeric comparison (age > 89) fails on this column (VARCHAR vs INTEGER), so the
-- string form below is required.
-- Mapping: '> 89' -> 90 for age filtering and descriptive statistics (section 2.2)
-- Exclusions: 530 stays with numeric age < 18 plus 95 stays with age unrecorded
-- (the latter cannot satisfy the adult predicate) = 625 excluded
-- Total: 200,859 -> 200,234 adult stays
CREATE OR REPLACE VIEW eicu_adult_stays AS
SELECT
    p.patientunitstayid,
    p.uniquepid,
    CASE WHEN p.age = '> 89' THEN 90
         ELSE TRY_CAST(p.age AS INTEGER) END AS age_mapped,
    p.gender,
    p.unitdischargestatus,
    p.hospitaldischargestatus,
    CASE WHEN p.hospitaldischargestatus = 'Expired' THEN 1 ELSE 0 END AS hospital_death
FROM eicu_patient p
WHERE p.age = '> 89' OR TRY_CAST(p.age AS INTEGER) >= 18;

-- Step 2: CRT records from nurseAssessment
-- cellattribute = 'Capillary Refill'
-- state values: 'normal', '< 2 seconds', '> 2 seconds'
-- site values: 'hands', 'feet'
-- Binary threshold: normal + < 2s = normal; > 2s = abnormal
CREATE OR REPLACE VIEW eicu_crt_records AS
SELECT
    na.patientunitstayid,
    na.nurseassessoffset,
    na.cellattribute,
    na.cellattributevalue,
    CASE
        WHEN na.cellattributevalue IN ('normal', '< 2 seconds') THEN 'normal'
        WHEN na.cellattributevalue = '> 2 seconds' THEN 'abnormal'
        ELSE NULL
    END AS binary_state,
    CASE
        WHEN na.cellattributevalue = 'hands' THEN 'hands'
        WHEN na.cellattributevalue = 'feet' THEN 'feet'
        ELSE NULL
    END AS site
FROM eicu_nurseAssessment na
WHERE na.celllabel LIKE '%apillary%';

-- Step 3: APACHE IV scores for selection bias analysis
CREATE OR REPLACE VIEW eicu_apache_scores AS
SELECT
    apr.patientunitstayid,
    apr.apachescore AS apache_iv_score,
    apr.apacheversion
FROM eicu_apachePatientResult apr
WHERE apr.apacheversion = 'IV';

-- Step 4: Export
COPY (
    SELECT * FROM eicu_adult_stays
) TO 'output/eicu_adult_stays.csv' (HEADER, DELIMITER ',');

COPY (
    SELECT * FROM eicu_crt_records
) TO 'output/eicu_crt_records.csv' (HEADER, DELIMITER ',');

COPY (
    SELECT * FROM eicu_apache_scores
) TO 'output/eicu_apache_scores.csv' (HEADER, DELIMITER ',');
```
