## Supplementary material for "Operational Heterogeneity of Capillary Refill Time Across Two Public ICU Databases: A Methodological Analysis": SQL queries and Python analysis scripts: verify_checksums.py

```
"""
verify_checksums.py — SHA256 verification for source data files
Usage:
    python verify_checksums.py --generate   # Generate checksums from local files
    python verify_checksums.py --check      # Verify files against checksums
"""
import hashlib
import os
import argparse
import glob

CHECKSUM_FILE = 'checksums_sha256.txt'

def sha256(filepath):
    h = hashlib.sha256()
    with open(filepath, 'rb') as f:
        for chunk in iter(lambda: f.read(8192), b''):
            h.update(chunk)
    return h.hexdigest()

def find_data_files(base_dirs):
    files = []
    for d in base_dirs:
        for ext in ['*.csv.gz', '*.csv', '*.parquet']:
            files.extend(glob.glob(os.path.join(d, '**', ext), recursive=True))
    return sorted(files)

def generate_checksums(data_dirs, base_dir='.'):
    lines = ['# SHA256 checksums for source data files\n']
    for f in find_data_files(data_dirs):
        rel = os.path.relpath(f, base_dir)
        cs = sha256(f)
        lines.append(f'{cs}  {rel}')
        print(f'  {cs}  {rel}')
    with open(CHECKSUM_FILE, 'w') as fh:
        fh.write('\n'.join(lines) + '\n')
    print(f'\nChecksums written to {CHECKSUM_FILE}')

def verify_checksums():
    if not os.path.exists(CHECKSUM_FILE):
        print(f'{CHECKSUM_FILE} not found. Run with --generate first.')
        return
    with open(CHECKSUM_FILE) as f:
        lines = [l.strip() for l in f if l.strip() and not l.startswith('#')]
    ok = 0
    fail = 0
    for line in lines:
        expected, relpath = line.split('  ', 1)
        if os.path.exists(relpath):
            actual = sha256(relpath)
            if actual == expected:
                ok += 1
            else:
                print(f'  FAIL: {relpath}')
                fail += 1
        else:
            print(f'  MISSING: {relpath}')
            fail += 1
    print(f'\nVerified: {ok} OK, {fail} failed/missing')

if __name__ == '__main__':
    parser = argparse.ArgumentParser()
    group = parser.add_mutually_exclusive_group(required=True)
    group.add_argument('--generate', action='store_true')
    group.add_argument('--check', action='store_true')
    parser.add_argument('--base-dir', default='.', help='Base directory for relative paths')
    parser.add_argument('--dirs', nargs='+', default=['data/raw/mimiciv_3_1', '/path/to/eicu_crd_2_0'])
    args = parser.parse_args()
    if args.generate:
        generate_checksums(args.dirs, args.base_dir)
    else:
        verify_checksums()
```
